# Feasibility and acceptability of contextually adapted AVATAR therapy for distressing voices in Ethiopia and India: a study protocol for the AVATAR3 study

**DOI:** 10.64898/2026.04.21.26348779

**Authors:** Thomas Ward, Atalay Alem, Thomas K. J. Craig, Koushik Sinha Deb, Surbala Devi, Abebaw Fekadu, Andrew Gumley, Charlotte Hanlon, Rebecca Kelly, Tsegahun Manyazewal, Eleni Misganaw, Isobel Murcutt, Edward Oshodi, Vaibhav Patil, Pratap Sharan, Yodit Tesfaye, Rohit Verma, Sophie Ul-Haq, Mar Rus-Calafell, Rashmi Choudhary, Medhanit Getachew, Amy Hardy, Mekdes Wondiye, Awoke Mihretu, Mamta Sood

## Abstract

**Introduction:** In many Low- and Middle-Income countries (LMIC), access to psychological therapies for psychosis remains extremely limited, contributing to significant treatment gaps and persistent inequalities in care. Novel interventions that are effective, scalable, and culturally acceptable across diverse settings are urgently needed. AVATAR therapy is an innovative digital intervention for distressing voices in psychosis, developed in the UK. The therapy enables voice-hearers to engage in a series of facilitated dialogues with a customized computer-based representation of their main distressing voice. AVATAR3 represents the first initiative to contextually adapt AVATAR therapy and evaluate its acceptability in two LMIC settings (Ethiopia and India).

**Methods and analysis:** We will establish Innovation and Implementation Hubs in Addis Ababa, Ethiopia (Centre for Innovative Drug Development and Therapeutic Trials for Africa (CDT-Africa) at Addis Ababa University (AAU) and Mental Health Service Users Association (MHSUA), Ethiopia) and New Delhi, India (All India Institute of Medical Sciences).

Phase 1 employs formative work and diverse stakeholder engagement to inform context-specific adaptations. Reflexive thematic analysis will be used, with data synthesis informed by the Cultural Adaptation of Scalable Psychological Interventions (CASPI) framework and Ecological Validity Model (EVM).

Phase 2 tests adapted AVATAR therapy through a parallel case series (n=15 per site, targeting 70% completion rate) measuring feasibility, acceptability, and safety indicators at baseline, 12-weeks, and 24-weeks. Qualitative research will explore the experiences of participants (n=10) and therapists (n=8) at each site.

**Ethics and dissemination:** Ethical approval has been obtained from Addis Ababa University College of Health Science Institutional Review Board, All India Institute of Medical Sciences (AIIMS) Institutional Review Board and the King’s College London (study sponsor) Research Ethics Committee. Findings will be disseminated to inform the implementation of AVATAR therapy across diverse international settings.

**Strengths and limitations of this study:**

- Interdisciplinary and participatory approach
- Contextual adaptation of a digital innovation
- Expert by experience leadership and involvement from the conception of the study
- The study will develop tools and share learning to support future digital mental health innovation across diverse international settings
- The case-series at each site will not have a control group

## Background

Voice-hearing has been documented across cultures and throughout human history (1). For some voice-hearers the experience is an enriching part of life. However, distressing voices feature across a range of mental health conditions (2) and are experienced by around 70% of those with a diagnosis of schizophrenia (3,4). The impact on individuals and families can be profound, with voices frequently persisting for years despite access to recommended pharmacotherapy (5). Psychological interventions such as Cognitive Behavioural Therapy for psychosis (CBTp) can reduce voice-related distress and facilitate recovery but are resource-intensive (6). Novel therapies that are effective, scalable, and culturally acceptable across diverse settings are urgently needed.

Emerging relational approaches (7) view voice-hearing as a fundamentally social experience, often involving communication with a characterized other (8,9). Distressing voices are typically experienced as powerful (even omnipotent) social agents, within relationships characterised by entrapment, disempowerment, and shame (10,11). AVATAR therapy (12) is a novel, digital therapy for distressing voices, which adopts a relational CBTp approach. The defining feature is the digital embodiment of the voice as an “avatar”, which is co-created with the voice-hearer. The therapist facilitates real-time dialogues allowing the person to experience dialogues “as if” communicating with the voice. Dialogues offer a space to explore personal meanings and to rescript disempowering interpersonal experiences, including discrimination and marginalization (13). The therapy aims to rebalance the relationship with the voice such that the person experiences increased empowerment and control (14).

AVATAR therapy has demonstrated robust efficacy in two fully powered UK-based randomised controlled trials (RCTs). In AVATAR1, the therapy delivered significant reductions in voice severity, with large effect sizes compared to an active control (15). AVATAR2, the subsequent multi-site RCT, tested two forms of the therapy: AVATAR-BRF (brief, 6 avatar dialogues) and AVATAR-EXT (extended, up to 12 dialogues), delivered across demographically diverse sites. AVATAR-BRF focused primarily on empowerment and building self-confidence, whilst AVATAR-EXT incorporated an additional phase aimed at developing a shared understanding of the voice grounded in the individual’s life history. Both treatment forms produced significant improvements in voice distress and severity at the end of therapy, with AVATAR-EXT also showing sustained benefits in a wide range of valued outcomes including voice frequency, empowerment, well-being, and recovery (16).

The UK-based trials have involved close collaboration with peer researchers and Experts by Experience (EbEs) and included large, diverse samples. Around 70% of participants recruited at the lead AVATAR2 site (South-East London), were from a racially marginalised group while the Glasgow site recruited a significant proportion of voice-hearers facing severe social deprivation, complex trauma, and intergenerational substance use. Working in these contexts has highlighted the profound impact of stigma, marginalisation, and social exclusion on voice-hearing. While significant advances have been made in standardising the AVATAR protocol, the powerful influence of social and cultural context remains fundamental to therapy personalisation (13). Following AVATAR2, a National Institute for Health and Care Excellence Early Value Assessment [NICE EVA] (17) recommended AVATAR therapy for deployment in the UK National Health Service to support real-world evidence generation.

In the UK, significant limitations remain in the availability of psychological therapy for distressing voices (18). This challenge is amplified in low- and middle-Income countries (LMIC^1^), where economic and systemic barriers contribute to significant treatment gaps and inequalities (20–22). The *Lancet Commission on Global Mental Health and Sustainable Development* (23) underlined the importance of training nonspecialists and use of digital technologies to deliver mental health interventions as a key driver of sustainable global change. Given that AVATAR therapy has demonstrated significant reductions in voice severity and distress within comparatively brief treatment durations (typically 8-10 sessions), it offers potential for scalable psychological intervention. However, while work in Germany (24) has led to inclusion in recent SL3 Guidelines (25) and replication efforts are underway in Denmark (26), Australia (27) and Canada (28), the therapy is yet to be tested in LMICs. Contextual adaptation is essential before implementation in new settings to ensure feasibility, acceptability, and effectiveness (29).

### Objectives

The AVATAR3 study will evaluate the feasibility and acceptability of contextually adapted AVATAR therapy within two distinct LMIC contexts: Ethiopia and India.

## Methods and analysis

### Study Design

The study will used mixed qualitative and quantitative methods across two phases. Phase 1 aims to contextually adapt AVATAR therapy for use in Ethiopia and India, guided by inclusive stakeholder engagement. Phase 2 aims to test the feasibility, acceptability, and safety of the adapted AVATAR therapy through a case series and qualitative research (See Figure 1). The project formally started on 1^st^ August 2024. Desk research commenced in January 2025. At the time of writing, formative work is complete at both sites, and qualitative research is ongoing. Phase 2 is scheduled to commence in October 2026.

**Figure 1.**
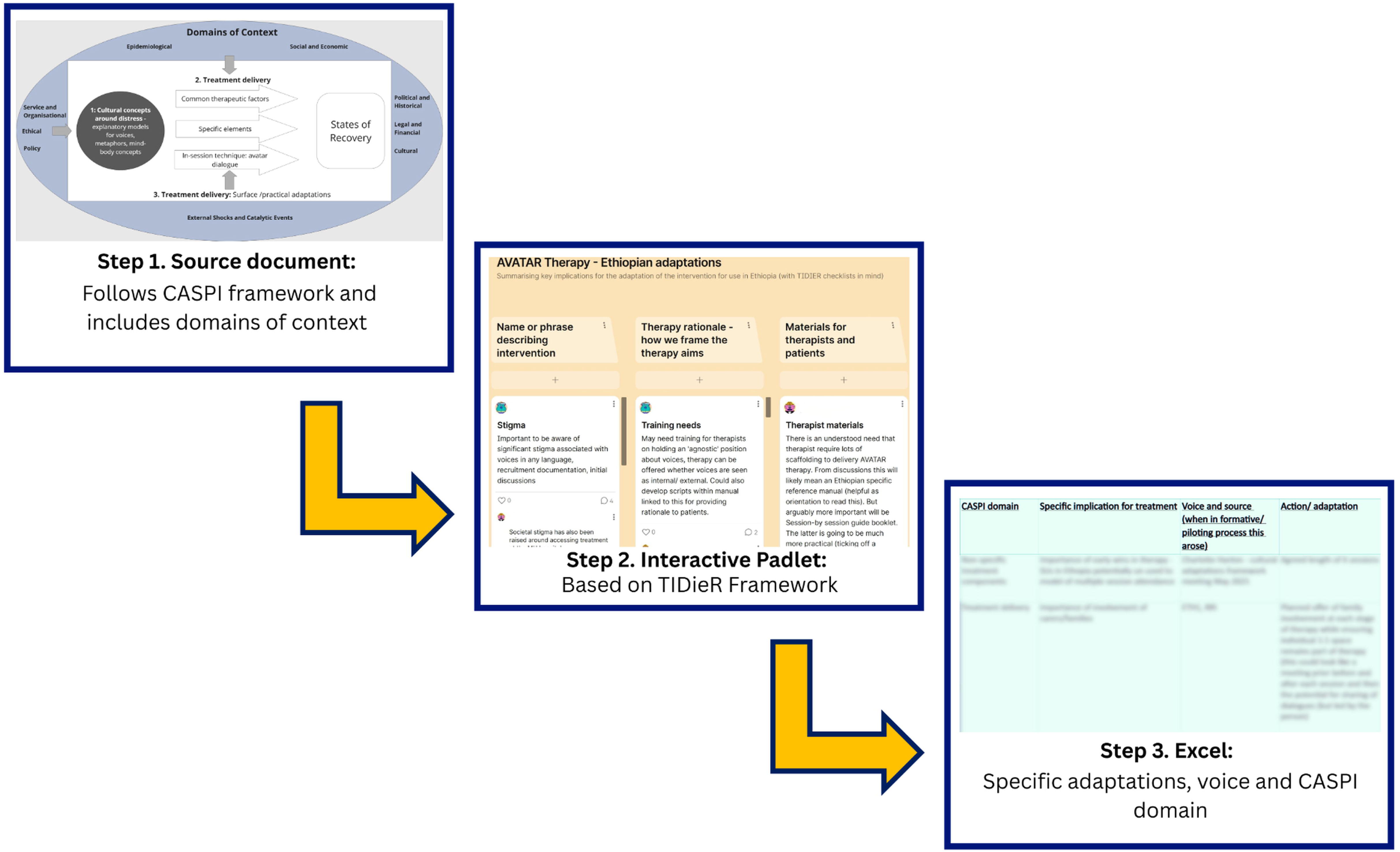
Illustration of the linkage between adaptation tools designed to facilitate impactful and inclusive stakeholder involvement.

#### Settings

Table 1 provides an overview of country and site characteristics. The sites are All India Institute of Medical Sciences (AIIMS), New Delhi, India and a partnership between Centre for Innovative Drug Development and Therapeutic Trials for Africa (CDT-Africa) at Addis Ababa University, and Mental Health Service Users Association (MHSUA) in Addis Ababa, Ethiopia. Each site contributes established clinical and research expertise and robust institutional infrastructure, which ensures equitable partnerships on the project and facilitates site-led innovation to support future implementation.

**Table 1.**
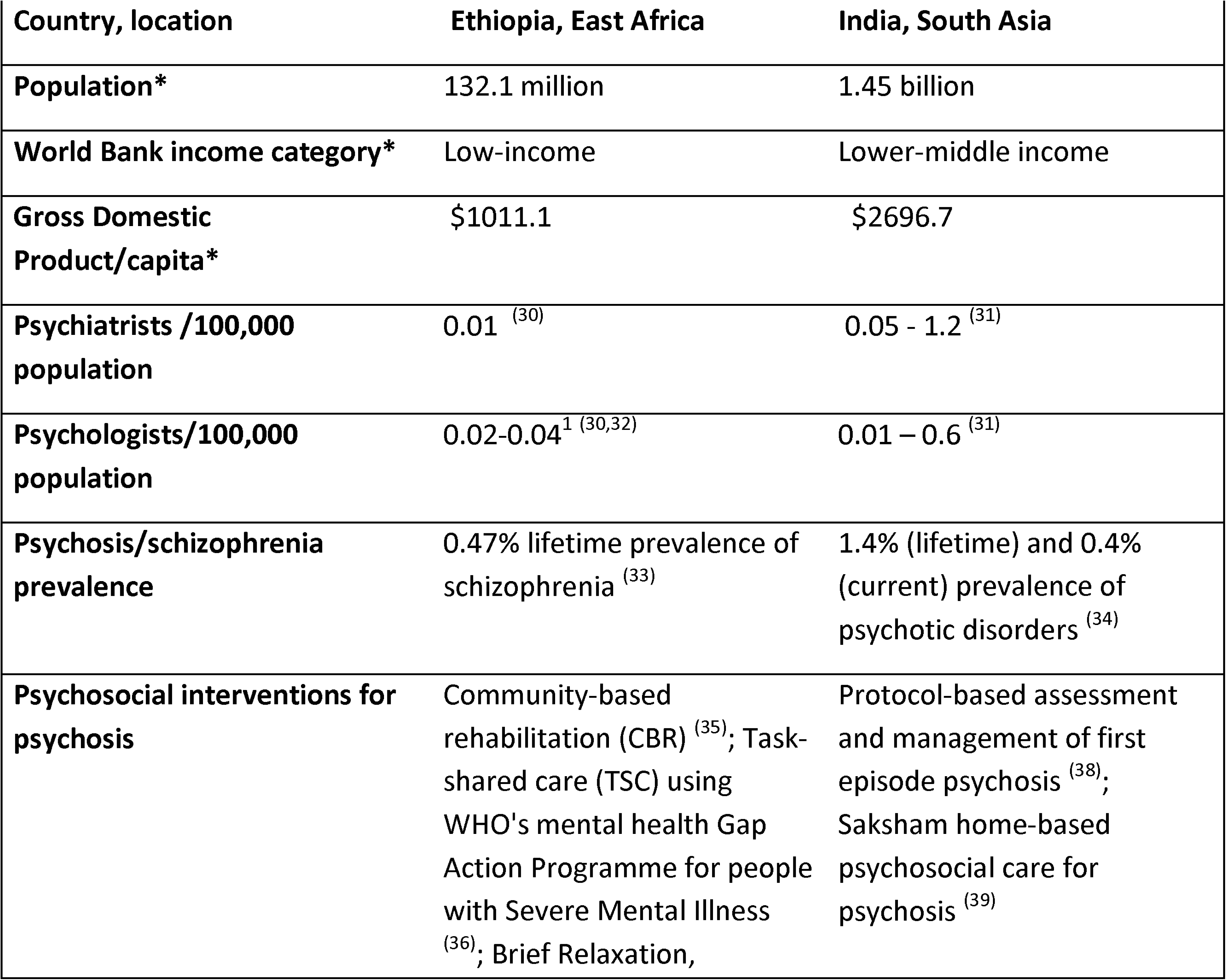
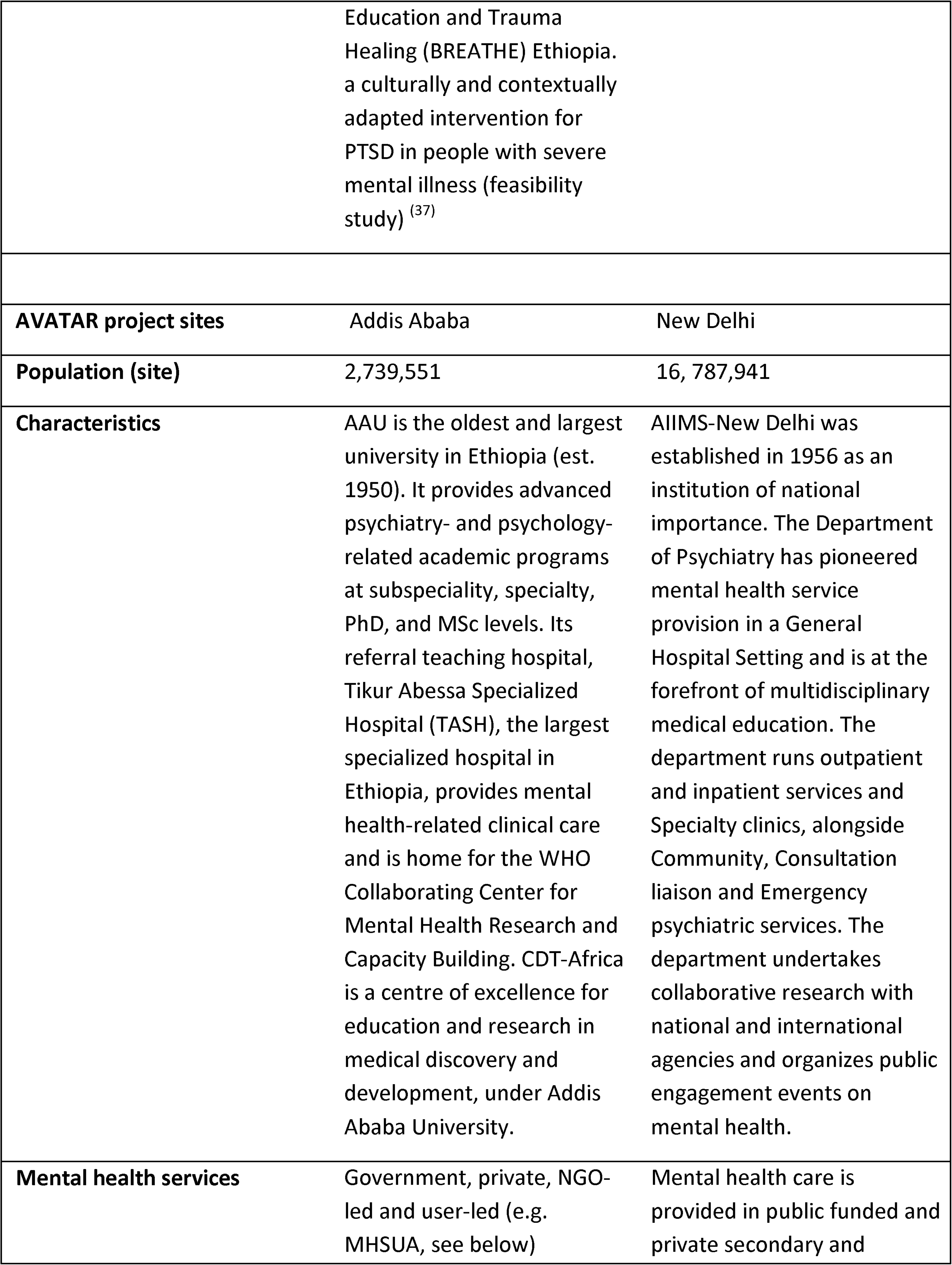
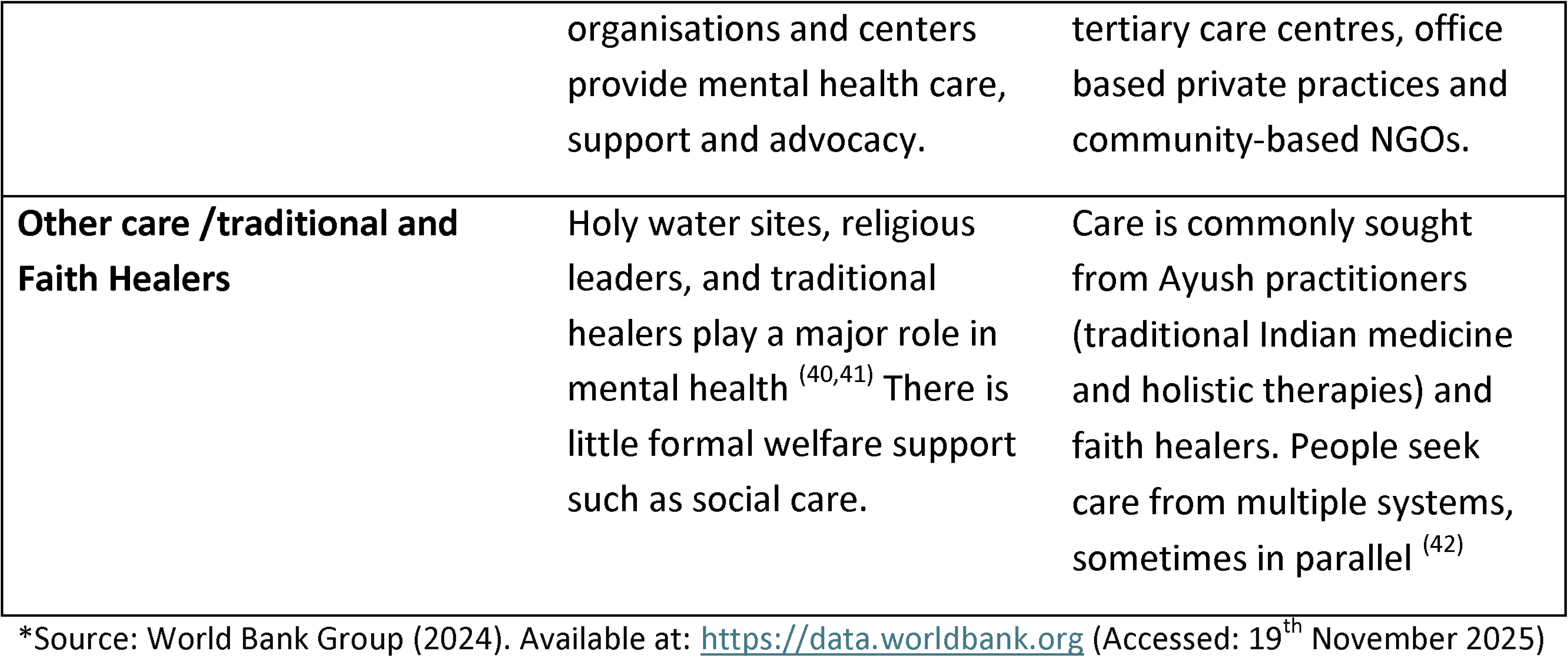
Characteristics of countries and project sites in AVATAR3.

### Contextualizing the problem: distressing voice-hearing

#### Prevalence and clinical need

Schizophrenia contributes more to the global burden of disease than any other mental health disorder (43), four-fold higher in LMICs compared to high-income regions (44), and with stigma identified as a key factor (45).

Schizophrenia and other psychoses represent a major health concern in Ethiopia and India. In Ethiopia, the large-scale rural (Butajira) community survey found a lifetime schizophrenia prevalence of 0.47% (33) with a treatment access gap of 41.8% (40). Significant investments have been made to upscale mental health service provision (32), but population coverage is low (46) and specialists remain scarce (47,48). For those able to access biomedical care, 71.7% do not receive minimally adequate care (40). Barriers to adherence include limited access to follow-up, lack of family support, and food scarcity when medications stimulate appetite (47,49,50). In India, epidemiological surveys have identified lifetime and current prevalence of psychotic disorders of 1.4% (around 19.6 million people) and 0.4% (around 5.6 million people) respectively, and a treatment gap of 75.5% (34), with significant regional disparities (51). In both countries, distressing voices in the context of psychosis are common, but opportunities to access effective psychosocial interventions are scarce.

#### Explanatory models and help-seeking

In Ethiopia, mental health difficulties are commonly attributed to supernatural or religious causes, or to drug or alcohol consumption. Societal stigma is common, with psychosis especially stigmatized (52). Qualitative research highlights significant voice-related distress, social exclusion and negative impact on sense of self-worth (53). Traditional and faith healers are a common first treatment choice (54).

In India, individuals experiencing a first episode of psychosis commonly attribute this to spiritual causes, most commonly “black magic”, with attributions to social, interpersonal, or biological factors relatively uncommon (55). Indian-led studies identify significant voice-related distress and impairment, with evidence of associations between beliefs about voices, response styles, and voice severity (56,57). Family perspectives influence help-seeking (58) with traditional healing centres (temples and dargahs) the first treatment choice for many (59). While the medical model of treatment for psychosis is dominant within mental health services, there is a growing interest in, and development of, psychological therapies for voice-hearers (60).

#### Theoretical framework and adaptation tools

We will follow the Adapting complex interventions to new Contexts (ADAPT) guidance (61). A range of data inputs will feed into the adaptation process (Figure 2). Data will first be synthesized within a Source Document based on the Cultural Adaptation of Scalable Psychological Interventions framework (CASPI) (62), which includes cultural concepts of distress, treatment components, treatment delivery, and states of recovery as potential domains for cultural adaptation (Figure 1: Step 1), and the Ecological Validity Model (EVM) (63), which identifies key areas of therapy adaptation (e.g. language, metaphors, aims/ rationale). Adapted TIDieR templates will be used iteratively (64) to track responsive adaptation. To maximise accessibility and engagement, these templates will be presented using interactive ‘padlets’ (Figure 1: Step 2). Once agreed by stakeholders, specific adaptations will be tracked alongside adaptation source (Figure 1: Step 3).

**Figure 2.**
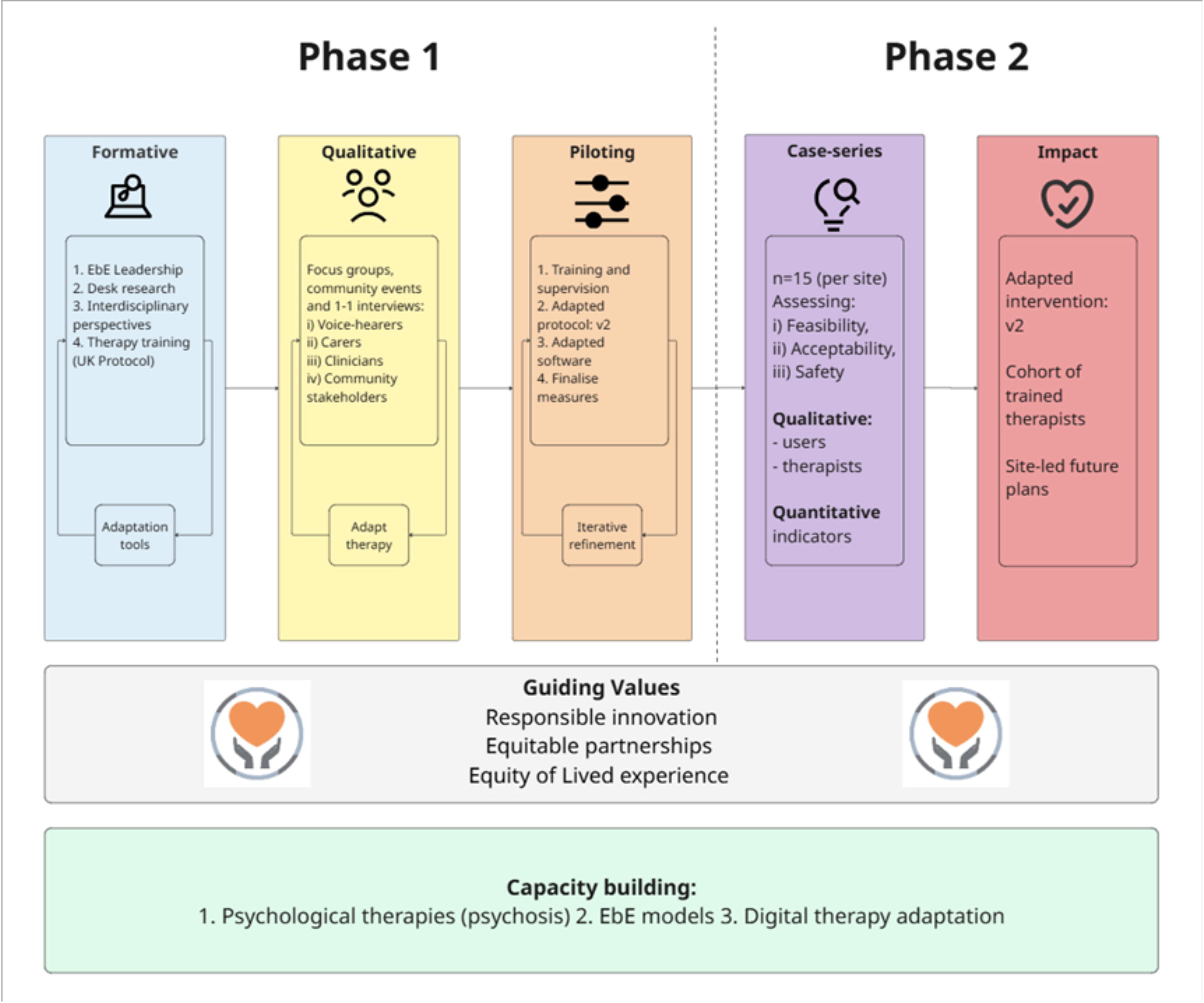
A study overview illustrating the two phases of the AVATAR3 project.

#### Hypothesized mechanisms of action

Phase 1 aims to ground the contextually adapted therapy within the site-specific context (62) while maintaining the following hypothesized mechanisms of action:

1. The digital embodiment of the voice (the personalized avatar) delivering a high sense of voice presence (65).
2. Anxiety reduction over the course of therapy.
3. Reductions in appraisals of voice power.
4. Increased appraisals of self-empowerment.

The adapted protocol and software will be shared with diverse stakeholders for iterative refinement prior to testing within the case series (Phase 2).

#### Patient and Public involvement (PPI)

Within our team, research partners MHSUA-Ethiopia have strongly advocated for use of the term Experts by Experience (EbEs) rather than PPI. As outlined in Phase 1 (below) EbE leadership and involvement has featured from the conception of the project and will continue across the duration of the study including dissemination and future planning.

### Phase 1

#### Formative Work

##### Experts by Experience (EbE) leadership and involvement

EbE leadership and involvement ensures that research remains applicable and relevant to real-world concerns and is conducted in a culturally respectful and accessible manner. Integrating lived experience is an ethical imperative, grounded in the principle that decisions should not be made without the meaningful participation of those most directly affected. The practice of incorporating lived experience will evolve through ongoing reflection and iterative learning, capturing, and building upon the knowledge generated (our Lived Experience framework is provided in Supplementary File 1). Sites will share learning and establish an involvement legacy, tailored to each context. A UK-based Lived Experience Lead (salaried member of the research team) will support EbE involvement across sites.

At AIIMS, the team will build on successful EbE involvement and co-design during a previous project, which developed a blended digital intervention (Saksham) for people with a diagnosis of schizophrenia (and caregivers) (66). The AIIMS team will recruit a Lived Experience Lead role who will receive training to co-deliver qualitative interviews and be involved in analysis. The team will also involve a wider EbE group (comprising individuals with lived experience of voice-hearing and carers) across diverse research activities.

In Ethiopia, EbE involvement is led by MHSUA-Ethiopia, a peer-led organisation representing people with lived experience of mental health difficulties that advocates for services, awareness, and human rights. MHSUA-Ethiopia has been involved from the inception of the project and played an important role in shaping the research proposal. While MHSUA has previously collaborated on a range of research projects, this study is the first time the organization has been formally contracted as a research partner. MHSUA holds a budget and has an MHSUA-employed Research Worker. This represents a crucial step in capacity building and equity in partnership. Ongoing MHSUA involvement includes: full participation at site meetings; an MHSUA representative leading the EbE discussion at the Responsible Research and Innovation (RRI) workshop (see Section 2); co-production of the adaptation tools, and co-facilitation of inclusive stakeholder engagement; contribution to therapy training; establishing the battery of outcome measures for the case series; participation at dissemination and community engagement events. Further opportunities for impactful involvement continue to emerge as the study proceeds.

##### 1. Desk research

Preliminary scoping work for each site focused on five key areas: cultural context, voice phenomenology, explanatory models, treatment approaches, and implications for AVATAR therapy. A summary including key conclusions and emerging questions can be found in Supplementary File 2. Context mapping considered cultural, social, and economic factors, as well as health system and service delivery considerations (see Supplementary File 3). To address likely gaps in research published in English, international partners shared quantitative and qualitative research and relevant publications in other languages.

##### 2. Interdisciplinary Engagement

In line with a commitment to ethical research, international partners, independent interdisciplinary experts, and diverse EbEs were brought together for a two-day RRI workshop, hosted by the UK team and facilitated by The Discovery Research Platform for Medical Humanities (Durham University). The workshop explored ethical and societal challenges relating to the study. It was shaped according to core principles of RRI, with a focus on inclusive dialogue and critical reflection about both positive impacts and potential unintended negative consequences of innovation (67). The methods and outputs of this workshop, which took place at the commencement of the adaptation work (March 2025), will be shared in a separate publication. Interdisciplinary engagement is intended to be iterative over the course of the study.

##### 3. Therapy training (UK protocol)

AVATAR “champions” (clinicians leading adaptation at each Hub) have been trained in the UK AVATAR therapy protocol. This represents an important early stage of the adaptation work, equipping partners with an understanding of the therapy methods and mechanisms of action. Training was conducted separately for each site through a combination of face-to-face training workshops and self-directed learning (reviewing therapy manual and therapy demonstrations). Content focused on core values and techniques associated with the therapeutic approach (e.g., therapy stance, engagement, assessment of voices). There was strong EbE involvement in the development and delivery of training. The Champions will lead on further training of therapists in the adapted therapy protocol, with ongoing support, training, and supervision from the UK team. This activity highlighted an opportunity for capacity building through additional training around CBTp approaches, delivered to clinicians beyond the study.

#### Qualitative research (Stakeholder engagement)

Through a series of focus groups, key-informant interviews, and community engagement events, we will explore diverse perspectives on the acceptability of AVATAR therapy and the adaptation required for successful implementation within the cultural and religious contexts. Topic guides (see Supplementary File 4) have been iteratively developed at each site, informed by the theoretical framework (see above). Engagement includes four main groups:

1. People with lived experience of distressing voices (“Service Users”), including individuals who have these experiences but do not use the language of “psychosis” or “hearing voices”.
2. Caregivers, defined as those with direct experience of providing care and support to a friend or family member who experiences distressing voices in the context of psychosis.
3. Mental health workers, defined as individuals in paid clinical roles with direct experience of providing clinical care to someone who experiences distressing voices in the context of psychosis.
4. Wider community representatives and stakeholders at a national level (e.g., faith or religious leaders, regional/ national mental health networks, and critical thinkers across a range of disciplines).

We will use reflexive thematic analysis (68) to inductively code data and utilise our adaptation tools (see Figure 1) to synthesise themes.

#### Piloting

##### Therapy

An adapted Therapy Protocol (Version 1) will be completed following synthesis of the findings from the Formative work and Stakeholder engagement. Site Therapy Leads will receive close supervision from UK team on the delivery of this protocol within early training cases. Qualitative feedback will inform iterative refinement of the protocol (leading to Version 2) and identify further required software adaptation. As part of the piloting stage and in collaboration with EbE leadership, the Phase 2 measures will be finalised to ensure they are tailored to each context.

##### Software Adaptation

The Avatar Therapy System has been certified with a class I UKCA mark by Avatar Therapy Ltd. Core software elements are voice enrolment (customising the voice of the avatar) and face enrolment (customising how the avatar looks). For voice enrolment, the therapist records their own voice, and the software manipulates that voice along dimensions of pitch, vocal tract size, spectral tilt and temporal roughness. Slider controls allow the person to hear different variations of the therapist’s voice until a good match to their distressing voice is achieved (Figure 3 Top Left). As this process is based on the therapist’s own voice, the main adaptation is to the language and content of the recording.

**Figure 3.**
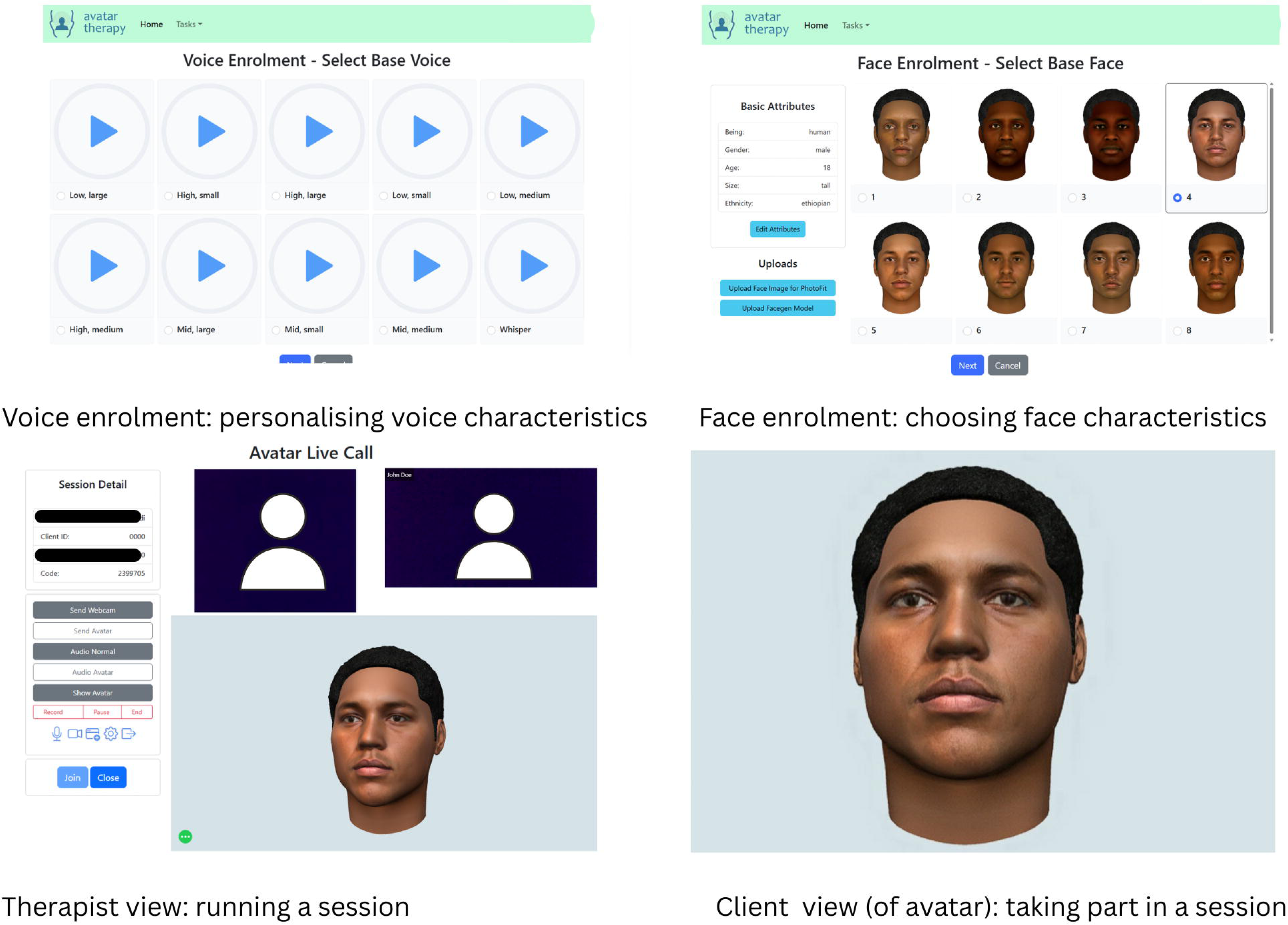
Illustration of Avatar software interface which is subject to ongoing adaptation for each site. *Avatar images shown are computer-generated representations created for illustrative purposes only and do not depict real individuals.

For face enrolment, the voice-hearer is presented with a gallery of faces (Figure 3 Top Right) that match their selection of basic attributes of the voice (e.g., age, gender, ethnicity). The person can customise facial shape, facial features, skin colour, skin texture, hair colour, and hairstyle. The standard software supports creation of bespoke avatars reflecting a range of ethnicities (current groupings include South Asian, Black African, White European, East Asian, Other). Through Phase 1 adaptation this standard creation process will be augmented to ensure the avatars are tailored to local diversity (including faces and cultural assets, e.g., hairstyles and headwear.)

### Phase 2

Phase 2 aims to test the feasibility, acceptability, and safety of the adapted AVATAR therapy (developed in Phase 1) via a case series and qualitative research at both the Ethiopian and Indian Hubs.

#### Data collection and analysis

With expected therapy completion rates of 70-75%, a sample of n=15 per site will result in (approximately) 11 therapy completers. Quantitative indicators of the process, acceptability, feasibility, and safety/harms associated with AVATAR therapy delivery will be collected. Therapy completion will be measured against a target of 70% (matching existing UK data). Safety will be assessed through monitoring of adverse events (including therapy relatedness). Preliminary indications of clinical impact will be assessed through data collected at baseline, 12-weeks, and 24-weeks follow-up.

Embedded qualitative research will be conducted at each site focusing on participant (n = 10 per site) and therapist (n = 8 per site) experiences of therapy. Data will be analysed using reflexive thematic analysis with adaptation tools used to contextualise themes.

At the culmination of Phase 2 we will return to the diverse stakeholders to share learning around implementation in context and the responsive adaptation.

Further information on Phase 1 and Phase 2 methods is available on OSF (69). Adapted therapy protocols and outcome measures will be piloted locally. The finalised study protocol and measures will be pre-registered prior to commencement of the case-series.

#### Outcomes

Based on study findings, the intention would be for each Hub to lead on plans for capacity building and potential scaling up access to the intervention, in partnership with the UK team.

## Discussion

The AVATAR3 study aims to test the feasibility, acceptability, and safety of contextually adapted AVATAR therapy in Ethiopia and India. The project will also build capacity for potential future scalability within site-led Innovation and Implementation Hubs. However, it is important to acknowledge that a study which brings an innovation from the Global North into India and Ethiopia (one of two African countries to never have been colonised) raises important questions. Potential objections could include cultural mistrust over what may be experienced as an unwelcome importing of western technology, or fear of a new form of colonialism through digital health research (see (70) for a thoughtful discussion of social psychiatry in the age of decolonisation). There may also be potential cultural dissonance with a therapeutic approach which has evolved within the philosophical and political context of the UK, where concepts relating to “the self” (e.g. self-esteem, self-empowerment, self-determination) often predominate and have been questioned with regards to implicit neoliberalism (71). Furthermore, when formulating a response to the global mental health crisis it is imperative to acknowledge the social and economic determinants of poor mental health (22). The impact of pervasive social inequalities is predicted to further escalate with the disproportionate impact of the climate crisis on LMIC. In this context, some might argue that investment in innovative digital health interventions may not be a priority when access to evidence-based non-digital mental health treatments remains scarce. However, the inherent risk in this approach is that LMICs become locked on a path of slow progress with an ever-widening gap to HIC, and opportunities for reciprocal learning and sharing of the benefits from emerging technologies are missed (72). Furthermore, digital approaches to healthcare delivery are receiving substantial governmental support in Ethiopia and India. The study will offer important opportunities for learning regarding the deployment of software as a medical device at the two sites. We are also developing adaptation tools that we hope will aid future digital mental health innovation across diverse international settings.

The AVATAR3 study involves a model of equitable collaboration between international partners, informed by a shift from a deficit-focused view of LMICs to one which engages with strengths, assets and opportunities for reciprocal learning (73). Crucially, this study does not position the Ethiopian and Indian Hubs as “test-sites” for an intervention which has achieved its final form in the Global North. Rather the study involves LMIC leadership early in innovative digital work, providing a platform to shape emerging interventions and facilitate effective and responsible implementation across a broad range of contexts. Phase 1 work has identified partner-led innovations including pioneering models of EbE involvement which will inform AVATAR therapy delivery in the Global North and other international settings. MHSUA-Ethiopia has identified the importance of establishing a sustainable legacy which includes (but is not limited to): improved accountability of research with respect to EbE involvement; development of models of peer supported interventions; establishing long-term funding of peer roles; and ensuring EbE perspectives are impactful at the policy level. To mitigate potential unintended consequences and deliver a positive sustainable legacy for the study we are committed to the following principles:

1. Equity in partnership.
2. Responsible Research and Innovation.
3. Equity of lived experience expertise.

Beyond AVATAR therapy, the broader ambition is to strengthen capacity for delivering psychological interventions for people with psychosis in Ethiopia and India.

## Data Availability

No datasets were generated or analysed as part of the current manuscript.

https://osf.io/zt4ra/files/d69jp

https://osf.io/zt4ra/files/3buem

https://osf.io/zt4ra/files/yrxgv

https://osf.io/zt4ra/files/mf4zu

## Ethics and dissemination

Ethical approval has been obtained from Addis Ababa University College of Health Science Institutional Review Board, All India Institute of Medical Sciences (AIIMS) Institutional Review Board and the King’s College London (study sponsor) Research Ethics Committee. Findings will be disseminated to inform the implementation of AVATAR therapy across diverse international settings.

## Acknowledgements

We acknowledge the support and expertise of collaborators Professor Angela Woods, Professor Ben Alderson-Day and Mary Robson at the Discovery Research Platform for Medical Humanities (The Platform | The Discovery Research Platform for Medical Humanities). AH and TW acknowledge funding from the Maudsley Biomedical Research Centre at South London and Maudsley NHS Foundation Trust and King’s College London. M.R.-C. discloses support for the publication of this work from the Sofja Kovalevskaja Award from the Alexander von Humbold Foundation and the Ministry of Education and Research in Germany (3.2-1210962-GBR-SKP). This study was supported by NHS Research Scotland through the Chief Scientist Office and the NHS Scotland Mental Health Network (A.G., no. NRSMHN/2021/01). CH, AA and EM receive support from the National Institute for Health and Care Research (NIHR) through the NIHR Global Health Research Group on Homelessness and Mental Health in Africa (NIHR134325) using UK aid from the UK Government. The views expressed in this publication are those of the authors and not necessarily those of the NIHR or the Department of Health and Social Care. CH, AA and EM receive funding support from Wellcome Trust through grant 222154/Z20/Z. CH also receives support from WT grant 223615/Z/21/Z. AA and AM receive funding from Developing Excellence in Leadership, Training and Science Initiative II (DELTAS Africa II) Grant a joint funding support from Wellcome Trust and UK’s Foreign, Commonwealth and Development Office (FCDO).

## Statement on use of Artificial Intelligence (AI)

AI-assisted tools were used in a limited capacity to assist with grammar and language editing. These tools were not used in the analysis, interpretation of data, or generation of study findings. All outputs were reviewed and verified by the authors, who retain full responsibility for the content of the manuscript.

## Ethical Standards

The authors assert that all procedures contributing to this work comply with the ethical standards of the relevant national and institutional committees on human experimentation and with the Helsinki Declaration of 1975, as revised in 2008.

## Author Contributions

Conceptualization of study: TW, AM, MS, AA, TC, KSD, SD, AF, AG, CH, TM, RK, EM, YT, VP, PS, RV, SUH, RK. Funding acquisition: TW, AM, MS, AA, TC, KSD, SD, AF, AG, CH, TM, EM, VP, PS, RV, AH. Writing – first draft: TW, RK, AM, MS, EO, IM, CH, YT, EM, SUH, TC. Writing – review and editing: TW, AM, MS, AA, TC, KSD, SD, AF, AG, CH, TM, RK, EM, YT, EO, IM, VP, PS, RV, SUH, MRC, RC, MG, AH, MW. All authors jointly decided to submit the manuscript for publication.

## Supplementary Materials

The supplementary material for this article can be found at: https://osf.io/zt4ra/files/osfstorage

## Footnotes

1 LMIC is widely used but can be problematic (19) - its use here reflects the limited access to psychosocial interventions common to both India and Ethiopia.

