## Supplementary File 4 for "Feasibility and acceptability of contextually adapted AVATAR therapy for distressing voices in Ethiopia and India: a study protocol for the AVATAR3 study"

**Interview Schedule for Exploring Service Users' Views on How AVATAR Therapy Should be Culturally Adapted to Fit Indian/Ethiopian Context**

### Service User Topic guide (Lived Experience Interviews)

Purpose: The following topic guide will be used to inform the 1:1 interview with people with lived experience of voices who have consented to take part. The guide identifies the areas to be explored during the interview. Suggested questions are presented (with available prompts). The interviewer will choose the phrasing of questions to ensure that they are accessible to the person being interviewed while covering the area which is being explored.

GUIDANCE NOTES WILL BE ADDED TO CONSIDER PAYING ATTENTION TO LANGUAGE (USE OF PERSON'S OWN WORDS), AND ADAPTATION OF LANGUAGE FOR ACCESIBILITY.

### Introduction and orientation to interview

This section should take 5 minutes to complete.

*Interviewer will introduce themselves and share the following rationale for the interview in keeping with the following:*

Thank you for agreeing to speak with me today. In the United Kingdom, the term "hearing voices" is often used to describe experiences where a person hears someone or something speaking to them that people around them cannot. Individuals may describe and understand these experiences in different ways. For some, the experiences are a positive part of their life. However, for others the experience starts to cause distress and gets in the way of the person living their life.

AVATAR therapy has been developed and tested in the United Kingdom and has been shown to be helpful for people who hear distressing voices. The purpose of the project here in <India/ Ethiopia> [DELETE AS APPLICABLE] is to hear different people's opinions about AVATAR therapy so we can understand how to adapt it to fit <India/ Ethiopia> [DELETE AS APPLICABLE].

We will start by exploring your understanding of your voices *[or alternative term tailored to individual and context]* and then move on to sharing some more information on what the therapy involves in the United Kingdom. We are interested to hear your thoughts about how the therapy fits for <India/ Ethiopia> [DELETE AS APPLICABLE] and any ways we can adapt the approach to make it fit better.

This interview will take approximately one hour to complete.

There are no right or wrong answers. We can take breaks at any point if helpful. The researchers will keep your responses confidential. We will make sure that you cannot be identified in any report. So, please express yourself freely.

If you have any questions at any point, please do not hesitate to ask.

*[Interviewer Guide: Review information sheet and consent form. Ask if they have any questions. Then move to signing consent form.]*

### Part 1 Exploration of service users' understanding of voices

This section should take 15 minutes to complete.

As previously mentioned, today is primarily about hearing your views on how to adapt AVATAR therapy to best fit the <Indian/ Ethiopian> [DELETE AS APPLICABLE] context but firstly, we would like to know more about your experience of hearing voices *[or tailored term]*. We are keen to understand how people describe and understand these experiences in their local context. This will help us understand more about how AVATAR therapy might be adapted to be most helpful for people in <India/ Ethiopia> [DELETE AS APPLICABLE]. We would like to hear about what the experiences mean to you, and how you respond to them. If there are any questions you would rather not answer, please let us know – we can skip them.

##### Questions

*[Interviewer Guide: Tailor the questions to make them understandable for the person; alternative ways to ask the question and follow ups are provided in the bullet points].*

1) **Can you tell me about your experience of hearing voices?**
   - How would you describe/ explain these experiences to someone close to you who does not know about them?
2) **What do you think causes the experiences/ where do they come from?**
   - How do you make sense of/ understand the experiences?
3) **When you have the experiences how do you respond to them?**
   - Are there things you have found more or less helpful in how you respond to the experiences?

*Optional prompts/follow-ups if relationship between interviewee and voices has not yet been discussed:*

- Would you say that you have a relationship with the voices? [If Yes] Can you tell me about this relationship?

Thank you for sharing those experiences.

### General Orientation

We are now going to talk through the main aspects of the therapy. We are interested in your views and any changes which might help for the approach to fit for <India/ Ethiopia> [DELETE AS APPLICABLE]. We will do this in three parts. The first is to get your thoughts on how the avatar is created. The second is to get your views on the way in which the person is supported to have a conversation with the avatar they have created. Once you know more about the therapy, we will finish by asking some questions to understand any general thoughts or suggestions you might have based on everything you have seen and heard.

### Part 2 Exploring views about the creation of the avatar

This section should take 15 minutes to complete.

The first session of AVATAR Therapy involves the person discussing their experience of hearing a voice with the therapist. The person then creates their own avatar. The person can make this avatar look and sound like the main voice they hear. The person might choose to create an avatar in human or non-human form.

The video you are about to watch shows the process of someone creating an avatar. We will check in after watching the video. I will then ask you some questions about the video you have just watched. If you need to stop the video at any point, please let me know.

***[Interviewer shares materials including still images and videos recorded in local language which have been created by each site team to show the avatar creation in an accessible form which is tailored to the context. These can be flexibly used and tailored to make accessible to the individual].***

**Questions:**

1) **What do you think about what you have seen (the avatar creation)?**
2) **How do you think you would find creating an avatar?**
   - What might help you (or someone else) to engage?
   - What might put you (or someone else) off?
3) **What kinds of avatars do you think people might create?**
   - What non-human forms might people create?
4) **Are there any ways in which we can improve this process (avatar creation) to help it to fit in the <Indian/ Ethiopian> [DELETE AS APPLICABLE] context**
   - Are there any choices that the software must have to be a good fit for <India/ Ethiopia> [DELETE AS APPLICABLE]?
   - Are there any things the software must not/ should not do to be a good fit?

#### Bridge

Now that we have shared an understanding of what the avatar is and how it is created, we will move on to how the avatar is used in the therapy. Before we go into these:

Do you have any questions or thoughts you want to share based on what we have just discussed?

### Part 2: Exploring views on conversations with avatars

This section should take 15 minutes to complete.

Once the person has created their avatar, they then have 6-10 sessions which involve conversations with the avatar, supported by a therapist. The therapist controls what the avatar says and how they act based on what the person has shared about their experience. The person learns to stand up to the voice and take control in a courageous and confident way. Over time, as the person stands up to the avatar, and gains more confidence and strength, the avatar will change, becoming less hostile and more friendly towards the person. The conversation is usually around 10 minutes of each 60-minute session. Some time is spent before the conversation to plan what the person wants to say. Afterwards the person has an opportunity to talk through how they found the conversation. The therapy aims to reduce distress and help people to live life on their own terms.  We hope that through the conversations with the avatar the person starts to feel more powerful, confident and in control when the voices are present.

The video you are about to watch is an example of someone having a conversation with an avatar. We will check in after watching the video. I will then ask you some questions about the video you have just watched. If you need to stop the video at any point, please let me know.

***[Interviewer shares materials including still images and videos recorded in local language which have been created by each site team to show the avatar creation in an accessible form which is tailored to the context. These can be flexibly used and tailored to make accessible to the individual].***

**Questions:**

1) **What do you think about the conversation between the person and the avatar?**
2) **How do you think you would find this?**
    - What might help you (or someone else) to engage? / What might put you (or someone else) off?
3) **What do you think people might want to get from engaging in the therapy?**
    - What goals or aims might people have?
4) **Are there any ways in which we can improve the avatar conversations to help it to fit in the <Indian/ Ethiopian> [DELETE AS APPLICABLE] context?**

### Bridge

Now that we have shared an understanding of AVATAR therapy, we are interested in hearing any final thoughts you might have. Before we go into these:

Do you have any questions of thoughts you want to share based on what we have just discussed? [Open question]

### Part 3: General thoughts

This section should take 10 minutes to complete

Ok thank you. Let's move into the final set of questions

**Questions:**

1) **What are your overall thoughts about how this approach might work in <India/ Ethiopia> [DELETE AS APPLICABLE]?**
2) **What language, words or ideas could be helpful in explaining AVATAR therapy (i.e. conversations and avatar creation)? Are there any words or language or ideas that might be less helpful?**
3) **Do you have any final thoughts on the most important changes to help the therapy fit in <India/ Ethiopia> [DELETE AS APPLICABLE]?**

### Ending (with thanks)

Thank you for taking part in this interview today. How have you found talking to us today? Has talking through your experiences brought up any difficult feelings? *[If so time and space is given for the person to discuss, and support is provided.]*

Please feel free to get in touch after the interview if you have any further questions – The contact details are [Insert].
