## Supplementary File 3 for "Feasibility and acceptability of contextually adapted AVATAR therapy for distressing voices in Ethiopia and India: a study protocol for the AVATAR3 study"

### Domains of Context

This document represents early desk-based scoping to inform contextual understanding for the cultural adaptation of AVATAR Therapy in India and Ethiopia. It reflects preliminary drafting by the UK team prior to partner consultation and does not represent our current or final position. Content has since been revised through collaborative review with partners in Ethiopia and India. We share this version for transparency and to document how our methods and assumptions evolved over time.

The following section provides an overview of the initial desk-based review that mapped key contextual domains relevant to adaptation. Desk reviews are commonly used to identify potential contextual influences on intervention design and delivery (Greene et al., 2017). The summary below outlines the early evidence gathered and how it was initially organised to guide subsequent collaborative inquiry.

**Epidemiological context**

The epidemiological context confirmed the high rates of distressing voice hearing and psychosis in India and Ethiopia, comparable to rates observed worldwide (Sagar et al., 2020; Kebede & Alem, 1999). Studies have also shown a higher prevalence in men (Kebede et al., 2003).

**Social and economic context**

There is a social and economic context marked by high levels of poverty and homelessness in both India and Ethiopia. In India, treatment costs for both traditional healers and psychiatric care are significant burdens for families (Chisholm et al., 2000). In Ethiopia caregivers have been found to have high burden relating to financial problems (often due to costs of treatment), fear for personal safety, stigma and problems with social life (Asher et al., 2015). In both countries, significant gender differences exist in rates of psychosis, with substantially more men than women recorded as affected (Teferra et al., 2012; Alem et al., 2009).

**Cultural context**

The cultural context summarised what is known about voice hearing in the respective countries, which identified high levels of stigma, multiple explanatory models for voices, and pluralistic healing approaches including medication, visits to faith healers and psychological care (Saravanan et al., 2007; Raguram et al., 2002; Srinivasan & Thara, 2001; Souraya et al., 2018); which may impact on openness to seeking psychological treatment and discussing voice content.

**Environmental context**

The environmental context highlights the differences between urban and rural populations and their access to health services in both India and Ethiopia. In India, rural areas, where two-thirds of the population lives, have disproportionately fewer mental health services and workforce (Mathur et al., 2024). Similarly, in Ethiopia a large portion of the population live in rural settings and difficult to access terrains making it more difficult to access the already scarce healthcare facilities. However, many community-based initiatives are being developed in conjunction with existing facility-based care to improve access to care (Asher et al., 2022).

**Service/ Organisational context**

The service and organisational context reflects a significant treatment gap, with little access to psychiatric and psychological support. In India, there are only roughly 0.3 psychiatrists per 100,000 people, well below the global average (Mathur et al., 2024). In Ethiopia, a survey conducted by the Ministry of Health reported that there are currently only around 70 psychiatrists working in Ethiopia (approximately 0.7/ 1,000,000 population) (Ethiopia Ministry of Health, 2012). Moreover, demand for health system-based care is low. Therefore, the Ethiopia mental health care plan focuses on community-based awareness-raising and case detection, with primary care workers (nurses and health officers) tasked with making the diagnosis, initiating treatment (including prescription of psychotropic medication) and providing continuing care, with monthly supervision from a psychiatric nurse (Hanlon, 2016).

**Ethical Context**

Concerning the ethical context, potential benefits and harms of the intervention must be considered. For example, in India, familial structures and collective decision-making may complicate informed consent and confidentiality (Trivedi & Tripathi, 2015). In Ethiopia a large portion of the population seek treatment and support from traditional and faith healers, therefore engagement with these stakeholders will be essential to understanding and respecting existing cultural and religious beliefs (Hanlon, 2016). Consequently, there may be a conflict of interest for traditional healers; there may be concern that encouraging use of medical or psychological treatments will reduce demand for their own services (Selamu et al., 2015).

**Policy context**

The policy context in India highlighted the differences in responsibility for healthcare between state and national governments, leading to fragmented implementation (Duffy & Kelly, 2019). In Ethiopia, demand for physical and mental health system-based care is low. Therefore, the Ethiopian Federal Ministry of Health strategy mandates explicitly the integration of mental healthcare into every primary care facility in the country to scale up mental health care (Fekadu et al, 2015; Hanlon, 2016; Ethiopia Ministry of Health, 2021).

**Legal and financial context**

The legal and financial context clarified commonalities and differences between the sites. The cost for service users to travel to appointments and afford treatment remains a major factor in both settings (Chisholm et al., 2007; Fekadu et al, 2015; Selamu et al., 2015). AIIMS represents a large institution recognised as an Institute of National importance by the Indian Government. This status brings opportunities for scale up through the affiliated AIIMS sites. These are distributed across India, guided by an institutional mission of offering equitable access to treatment across the underserved communities of India. At AIIMS there are multi-level governance processes, including governmental (Health ministry) approvals required to undertake the project. AAU in Ethiopia and the affiliated CDT-Africa, are well placed in a strategic sense for potential roll out beyond the project- however shortage of financial resources represents a significant barrier. Emerging legislation in areas such as data protection and regulation of medical devices are also relevant to work on the project.

The political and historical context may impact the countries differently. Ethiopia is one of two African countries that were not colonised and may therefore be more resistant to externally developed 'Western' interventions (Wondie & Abawa, 2019). The historical links between UK and India mean significant interaction and cultural exchange has taken place; however colonial legacies may influence perceptions of interventions developed in UK. The concept of avatar embodiment is Indian in origin (and an influence on the development of AVATAR therapy) and this may facilitate connection with the therapeutic use of avatars within the Indian content (although there remains a question as to whether the digital aspect could be a barrier for some).

**External shocks and catalytic events**

Finally, external shocks and catalytic events. In India, the COVID-19 pandemic disrupted services and accelerated digital initiatives such as [Tele Manas](https://telemanas.mohfw.gov.in/home) services but also widened existing disparities in access to care. In Ethiopia, two episodes of inter-ethnic violence in September and November 2018 in Meskan and Mareko districts caused at least 24 deaths, 167 injuries, and the displacement of thousands of people (Addis Standard, 2018). Conflict has also arisen between government forces and the Tigray people’s liberation front in November 2020 (BBC News, 2021), with continuing impacts on northern Ethiopia (Keynejad et al., 2024). With respect to India, risk of conflict in Kashmir is heightened in the light of a recent terrorist incident (BBC News 2025). This occurs amidst the background of reported tensions within the country between India’s diverse communities, in particular between Hindu and Muslim communities rooted in a history of partition and associated with an increased influence of Hindu Nationalism within Government.

BBC News. (2021, November 4). *Ethiopia: Tigray conflict continues amid humanitarian crisis.* Retrieved from: [www.bbc.com/news](https://www.bbc.com/news)

BBC News. (2025, March 2). *Kashmir tensions rise after recent attacks.* Retrieved from: [www.bbc.com/news](https://www.bbc.com/news)

Chisholm, D., James, S., Sekar, K., Kumar, K. K., Murthy, R. S., Saeed, K., & Mubbashar, M. (2000). Integration of mental health care into primary care: Demonstration cost–outcome study in India and Pakistan. *British Journal of Psychiatry, 176*(6), 581–588. doi:10.1192/bjp.176.6.581

Duffy, R. M., & Kelly, B. D. (2019). India’s Mental Healthcare Act, 2017: Content, context, controversy. *International Journal of Law and Psychiatry, 62*, 169–178. doi:10.1016/j.ijlp.2018.08.002

Ethiopia Ministry of Health. (2012). *Mental health workforce report.* Ministry of Health, Addis Ababa. Retrieved from: [www.moh.gov.et](https://www.moh.gov.et)

Ethiopia Ministry of Health. (2021). *Health Sector Transformation Plan II (HSTP II) 2020/21–2024/25.* Ministry of Health, Addis Ababa. Retrieved from: [www.moh.gov.et](https://www.moh.gov.et)

Fekadu, A., & Thornicroft, G. (2014). Global mental health: Perspectives from Ethiopia. *Global Health Action, 7*(1), 25447. doi:10.3402/gha.v7.25447

Greene, M. C., Jordans, M. J. D., Kohrt, B. A., Rahman, A., & Patel, V. (2017). Addressing contextual factors in intervention design and implementation for mental health and psychosocial well-being in low-resource settings. *Global Mental Health, 4*, e11. doi:10.1017/gmh.2017.10

Hanlon, C. (2017). Next steps for meeting the needs of people with severe mental illness in low- and middle-income countries. *Epidemiology and Psychiatric Sciences, 26*(4), 348–354. doi:10.1017/S2045796016001013

Heim, E., & Kohrt, B. A. (2019). Cultural adaptation of scalable psychological interventions: A new conceptual framework. *Clinical Psychology in Europe, 1*(4). doi:10.32872/cpe.v1i4.37679

Hoffmann, T. C., Glasziou, P. P., Boutron, I., Milne, R., Perera, R., Moher, D., Altman, D. G., Barbour, V., Macdonald, H., Johnston, M., Lamb, S. E., Dixon-Woods, M., McCulloch, P., Wyatt, J. C., Chan, A.-W., & Michie, S. (2014). Better reporting of interventions: Template for intervention description and replication (TIDieR) checklist and guide. *BMJ, 348*, g1687. doi:10.1136/bmj.g1687

Kebede, D., & Alem, A. (1999). Major mental disorders in Addis Ababa, Ethiopia. I. Schizophrenia, schizoaffective and cognitive disorders. *Acta Psychiatrica Scandinavica, 100*(S397), 11–17. doi:10.1111/j.1600-0447.1999.tb10688.x

Mathur, R., Chawla, N., & Chadda, R. K. (2024). Mental health services in rural India: A big challenge still to be met. *BJPsych International, 21*(4), 93–96. doi:10.1192/bji.2024.25

Raguram, R., Venkateswaran, A., Ramakrishna, J., & Weiss, M. G. (2002). Traditional community resources for mental health: A report of temple healing from India. *BMJ, 325*(7354), 38–40.

Sagar, R., Dandona, R., Gururaj, G., Dhaliwal, R. S., Singh, A., Ferrari, A., Dua, T., Ganguli, A., Varghese, M., Chakma, J. K., Kumar, G. A., Shaji, K. S., Ambekar, A., Rangaswamy, T., Vijayakumar, L., Agarwal, V., Krishnankutty, R. P., Bhatia, R., Charlson, F., … Dandona, L. (2020). The burden of mental disorders across the states of India: The Global Burden of Disease Study 1990–2017. *The Lancet Psychiatry, 7*(2), 148–161. doi:10.1016/S2215-0366(19)30475-4

Saravanan, B., Jacob, K. S., Johnson, S., Prince, M., Bhugra, D., & David, A. S. (2007). Belief models in first-episode schizophrenia in South India. *Social Psychiatry and Psychiatric Epidemiology, 42*(6), 446–451. doi:10.1007/s00127-007-0186-z

Selamu, M., Asher, L., Hanlon, C., Medhin, G., Hailemariam, M., Patel, V., Thornicroft, G., & Fekadu, A. (2015). Beyond the biomedical: Community resources for mental health care in rural Ethiopia. *PLOS ONE, 10*(5), e0126666. doi:10.1371/journal.pone.0126666

Souraya, S., Hanlon, C., & Asher, L. (2018). Involvement of people with schizophrenia in decision-making in rural Ethiopia: A qualitative study. *Globalization and Health, 14*(1), 85. doi:10.1186/s12992-018-0403-4

Thara, R., & Srinivasan, T. N. (2000). How stigmatising is schizophrenia in India? *International Journal of Social Psychiatry, 46*(2), 135–141. doi:10.1177/002076400004600206

Wondie, Y., & Abawa, M. (2019). Westernization versus indigenization in the context of global mental health: Training and services in Ethiopia – University of Gondar in focus. *International Journal of Mental Health, 48*(4), 257–271. doi:10.1080/00207411.2019.1644139
