## Supplementary File 2 for "Feasibility and acceptability of contextually adapted AVATAR therapy for distressing voices in Ethiopia and India: a study protocol for the AVATAR3 study"

**Voice Hearing in India and Ethiopia: Desk Research**

This document represents early desk-based research conducted using English-language literature to inform preliminary understanding of context for the cultural adaptation of AVATAR Therapy. It reflects initial drafting and early-stage thinking by the UK team prior to partner consultation. The content has since been revised and updated through collaborative review with project partners in Ethiopia and India. This version is shared for transparency and to document the evolution of our adaptation methods; it does not represent the final or current position of the team.

**Introduction**

Phase 1 of the AVATAR3 study aims to explore how best to adapt AVATAR Therapy in an ethically responsible and culturally acceptable manner to fit the Ethiopian and Indian context. Drawing on CBT adaptation methods, best practice guidelines recommend the process of adaptation begin with the gathering of information from the literature in relation to the cultures in question (Naeem et al., 2019). Therefore, the following working document aims to briefly summarise and synthesise the extant literature. This review is a working document, and a first step towards highlighting some of the potential issues and questions that might impact on the adaptation of AVATAR therapy for voice-hearers in India and Ethiopia. Further information and perspectives will be gathered at the RRI workshop in the UK, and from the subsequent stakeholder engagement work at each site.

In this review, we will briefly consider available evidence in both India and Ethiopia relating to the phenomenological experience of hearing voices, professional and lay perspectives and explanatory models, and the availability and acceptability of existing therapeutic approaches. The aim is to identify important questions that we hope the stakeholder engagement at each setting will help us to answer, so that we can best adapt AVATAR therapy for individuals experiencing voices in these countries.

**Voice hearing across cultures**

Hearing voices is often understood as a symptom of psychosis and can be viewed as requiring psychiatric or psychological interventions. However, how voice hearing is experienced, understood, and managed varies widely across cultural and socioeconomic contexts (Luhrmann et al., 2015b), and beliefs about the causes of mental illness can influence the choice of treatment and help-seeking pathways (Girma et al., 2024). It has been argued that modern psychosocial interventions developed in the West are underpinned by Western cultural values (Naeem et al., 2019), and this might influence the acceptability and effectiveness of these interventions in other cultures and contexts. For example, in contrast to Western societies, in which the idea of the independent self dominates, Eastern societies are reported to view the self as interdependent, characterized centrally by relational attributes, such as a familial self and a spiritual self (Kitayama & Salvador, 2024; Mihretu, 2021). It is important that these cultural factors are taken into consideration and explored prior to adapting and delivering psychosocial interventions developed in the West in non-Western countries (Naeem et al., 2019). The availability of psychological interventions for psychosis remains limited, particularly in low- and middle-income countries (LMICs)^[1]^, where economic and systemic challenges contribute to a significant treatment gap (Kaur & Pathak, 2017). Resource constraints, cultural beliefs, and explanatory models of mental health play an important role in shaping how voice-hearing is perceived—whether as a condition requiring medical treatment, a spiritual experience, or a social and familial issue (Luhrmann et al., 2015a), and this in turn impacts perceptions of psychosocial interventions and their relevance and acceptability.

**India**

In India, diverse cultural, religious, and social frameworks influence individual and collective understandings of voice-hearing, impacting methods of seeking support and treatment pathways (Luhrmann et al., 2015b). Additionally, the phenomenology of voice hearing may differ from that of high-income countries (HIC) like the UK, raising important questions about the acceptability of psychological interventions like AVATAR Therapy. This summary explores the cultural, explanatory, and treatment-related dimensions of voice hearing in India, exploring how voices are experienced, the models used to explain psychosis, and the treatment options available. By mapping out these factors, we can highlight key considerations for adapting AVATAR Therapy to the Indian context.

**Cultural context**

India is home to one of the largest populations in the world, with a rich diversity of languages, cultural practices, and environments, ranging from densely populated urban areas to more remote rural communities. Indian culture is deeply influenced by values surrounding family, religion, and spirituality, with a strong emphasis on communal ties. India is a country with a rich and diverse religious landscape, with Hinduism being the largest religion, followed by Islam, Christianity, Sikhism, Buddhism, and Jainism, among others.

An often-stated distinction between India and the UK lies in each society's individualistic versus collectivist perspectives. In HICs like the UK, individuals tend to view themselves as independent, self-reliant beings whose personal identity is largely autonomous and distinct from others (Geertz, 2020). In contrast, in cultures such as India, this is commonly framed in more relational terms, with identity being closely linked to family, community, and social connections (Neisser & Jopling, 1997). In more collectivist societies, personal identity is often understood in terms of one's role within a broader social circle, which influences the interpretation of mental health issues and the approach to seeking treatment (Khemani et al., 2020). For example, people who hear voices in India are more likely to stay with their families, who are seen as more supportive than in HIC, viewing the voices as a collective problem to solve (Luhrmann & Marrow, 2016). Decisions about seeking care are often guided by an individual's personal beliefs and the influence of family and friends (Khemani et al., 2020). Local cultural beliefs are not fixed but are multifaceted, and constantly evolving, highlighting the need for further research to understand their influence on clinical practice (Saravanan et al., 2007).

The impact of culture on the experience of voice hearing is a critical yet under-explored area of research (Ghanem et al., 2023). Early research assumed that voice-hearing impacts individuals similarly; however, more recent studies highlight that the phenomenology of voice-hearing can be shaped by the cultural context in which a person exists (Luhrmann et al., 2015b). In India and other potentially more collectivist cultures, some individuals may interpret voice hearing as spiritual, relational, or even protective (Luhrmann et al., 2015a). While cultural frameworks in HICs and LMICs can influence voice hearing experiences differently, these distinctions are not absolute. The UK, and particularly South-East London (one of the places where AVATAR therapy has been tested) is highly diverse, and some voice-hearers’ interpretations have aligned with those observed in India. This cultural variability emphasises the need to account for local contexts when considering the adaptation of AVATAR therapy across diverse cultural settings, while also highlighting that voice-hearing experiences can share commonalities across cultures, making it important not to assume difference where commonalities may exist.

**Voice phenomenology**

Phenomenology is a subjective experience of voices, encompassing their sensory qualities, content, and affect. Voices may be perceived as originating internally or externally, and they can range from benign, guiding, and even comforting to distressing, commanding, or persecutory (McCarthy-Jones et al., 2014). In India, some people's experience of hearing voices has been described as internal, softer, and conversational rather than intrusive, like they are more commonly seen in HICs (Luhrmann et al., 2015a). In a study by Luhrmann and colleagues (2015), they found that the voices in Chennai often represented common familial dynamics, for example, guiding the person while also scolding them, which they did not necessarily find threatening. At the same time, while some viewed their voices as 'fun' or positive experiences, some people understandably found hearing traditionally 'negative content' such as instructions to hurt themselves or others as distressing. In Chennai, it was common to hear voices discussing sex, and participants often felt intense shame around this type of voice content (Lebovitz et al., 2021). The experience of voices delivering taboo content is particularly relevant to AVATAR therapy, as it may influence how individuals engage with the intervention and the kinds of dialogues that emerge during therapy.

Luhrmann et al. (2015a) found that voice-hearers in their California sample were more likely to experience voices as a disrupted relationship between thought and the mind, perceiving them as intrusive and unreal. In contrast, in South India, voices were more often described as providing useful guidance. However, the perception of voices is not uniform within any setting. Some individuals in India accept their voices as part of their identity, particularly when linked to spiritual beliefs, while others experience significant distress when voices are associated with 'black magic,' affecting their self-perception, family dynamics, and treatment-seeking behaviour (Thara & Srinivasan, 2000). Importantly, many UK voice-hearers also understand their voices as external in origin, highlighting the need to avoid overly rigid cultural distinctions when considering the phenomenology of voice-hearing.

**Explanatory models**

In India, there are multiple explanatory models of psychosis, including spiritual factors, biomedical models, and traumatic events. Some families attribute psychosis to supernatural causes such as 'evil spirits' or 'black magic' (Raguram et al., 2002; Thirthalli et al.,2016). In a study of 131 people in South India, 70% of participants considered spiritual and mystical factors as the cause of their voices, for example in one focus group a man described their understanding of the voices as saying, ‘Most of us villagers go to the forest and work late nights alone, we get in contact with so many things there, you feel it directly. It’s because of that we think and believe this could be because of black magic.’ (Saravanan et al., 2007). The biomedical model is gaining recognition in India, particularly in urban areas with more access to education (Khemani et al., 2020). This is mainly due to exposure to media awareness campaigns and greater access to mental health services, which have contributed to this shift in understanding. Social and familial models also play a significant role in shaping how psychosis is understood in India. Some communities interpret voice hearing as a result of interpersonal conflicts emphasising the collective nature of the Indian culture (Johnson et al., 2012). There are many explanatory models of voice-hearing in India, and it is important that they not be dismissed or invalidated.

**Treatment approaches**

The cultural context in India impacts how people seek out treatment. Visiting traditional healing centres is the first treatment choice for many people in India, who seek help through visits to temples and dargahs (shrines or tombs of religious figures) (Raguram et al., 2002). Research conducted in Gwalior and Jaipur found that 69% and 40% of individuals, respectively, initially sought help from traditional healers. For some people in India, there is a stigma attached to seeking help from medical professionals. For example, some people feel that accepting medication means admitting that there is a mental health difficulty (Goyal et al., 2022). In a culture where community is important as well as status in relation to marriage, people can be reluctant to seek out professional help as they may label themselves as 'crazy' after seeking medical help, which could potentially make them 'less desirable' as a partner (Sinha & Ranganathan, 2020). The medical model of treatment for psychosis is prevalent in India, with the development of Early Intervention Services for Psychosis and medication being the primary form of treatment for voice-hearers (Keshavan et al., 2010). This being said, there is a growing development of psychological therapies for voice-hearers, including CBT-P, and a focus on using mindfulness to manage voices (Kumar, 2018). Many voice-hearers take a multifaceted approach to seeking out help for their symptoms, for example, using both traditional methods of faith healing as well as taking medication.

**Implications for AVATAR Therapy**

It is clear that there are important differences in the way that psychosis is understood, experienced, and treated in India in comparison with the UK. Cultural and religious contexts influence how people in India understand their voices, as many do not always view them as intrusive as many people in the UK do. Some find their voices guiding and helpful and seemingly mimic familial dynamics, highlighting the importance of a community in India however, some hear critical and scolding voices. Many voice-hearers also have a relationship with their voice, which could be a helpful starting point during AVATAR therapy, given the relational emphasis of the therapy. Considering the systemic implications of someone attending therapy could be important, as families play a significant role in treatment. Therefore, it is important to consider what the person is comfortable with and how much the broader support system is involved. Another important consideration is the multifaceted nature of treatment in India currently; it will be important to consider any other treatment the person may be receiving and use that to empower the person, aligning with their beliefs. It will be important to consider the gender, age, and educational level of those partaking in AVATAR therapy and how this may change their understanding of the therapy and trust in mental health services.

**Conclusions and questions**

Key questions for the stakeholder research and for subsequent adaptation include:

- Are there cultural/religious factors that might influence therapy engagement (e.g., perceived spiritual risks of engaging with voices)?
- Would voice-hearers prefer an adapted version of AVATAR Therapy that includes family involvement or integrates traditional healing perspectives?
- How do familial dynamics impact ability to engage with the therapy (i.e. speaking with an AVATAR of a family member)?
- Do voice-hearers who hear familial voices as scolding and/or guiding find this distressing? How many people who seek treatment have this understanding?
- What are the various understandings of “black magic” and how would this impact engagement with the therapy?

**Ethiopia**

**Cultural context**

Ethiopia has a large and diverse population with various ethnic groups, including but not limited to the Oromo, Amhara, Dawro, Keffa and Yem peoples (Alem et al., 1999; Girma et al., 2024). Amharic and Afaan Oromo are reported to be the most widely spoken languages. A fifth of the population are reported to live in urban settings, with the majority living in rural communities (Crummy & Marcus, 2025). Ethiopian society is largely religious (Mihretu, 2021) and as of 2012, census data indicated that 43% of the population were said to be affiliated with Orthodox Christianity, 34% with Islam, 19% with Catholicism and a further 1.5% with traditional beliefs (Crummy & Marcus, 2025). It has been proposed that those living in Ethiopia commonly attribute mental illness to supernatural causes (Girma et al., 2024; Mulatu, 1999). This context differs from locations where AVATAR therapy has been trialled to date. Exploring how voices are experienced by different ethnic groups, religious groups and, between those living in urban versus rural settings will be crucial to the adaptation of AVATAR Therapy in a culturally acceptable manner.

AVATAR therapy draws on a relational approach to understanding voices and works to understand the interpersonal relationship between voice-hearer and the main distressing voice (Rus-Calafell et al., 2020; Ward et al., 2020), with therapeutic targets focusing on helping the voice-hearer develop an increased sense of power and control, consistent with broader cognitive approaches (Ward et al., 2020). With this in mind, it will be important to understand how individuals within the different subgroups of Ethiopia relate to voices, particularly whether voices are seen as persecutory or distressing, as in AVATAR Therapy, the focus is always on what is distressing and interfering with life (Ward et al., 2020). This also leads to questions about the acceptability of AVATAR therapy as an intervention and how differences between groups may impact this.

**Voice phenomenology**

AVATAR therapy allows for face-to-face dialogue between the voice hearer and a computerised representation of the main voice (Ward et al., 2020), therefore, it will be important to gain a sense of how voices might be embodied in an Ethiopian context, which shall no doubt inform the software development side of the avatar creation process. For example, in a UK context, 69% of respondents in an analysis of a new phenomenological survey reported their voice(s) had person-like characteristics, such as gender, age, patterned emotional responses, or intentions, whereas only 16% of respondents reported their voice(s) as supernatural entities (Woods et al., 2015). We were not able to identify research published in English to date to indicate whether these findings from phenomenological studies apply in an Ethiopian context. After a thorough search of the literature, there appears to be a dearth of published work in the English language regarding the phenomenology of voice hearing in Ethiopia. One possible reason for the lack of published work on the phenomena of voice hearing in Ethiopia may be due in part to stigma attached to mental health conditions, creating a barrier to the access of treatment and subsequently exploration from a lived experience perspective (Girma et al., 2024). Thus, it appears much of the emphasis in Ethiopia has been placed on destigmatising mental health conditions, raising community awareness and expanding access to mental healthcare in the local community (Girma et al., 2024). It is also likely that research has been conducted into the phenomenological experience of voice-hearing that has not yet been published or translated into the English language. Stakeholder engagement work will add to our understanding of what is already known about voice phenomenology in Ethiopia.

**Explanatory models**

In Ethiopian traditional societies, it is reported that well-being is first and foremost secured and maintained through a peaceful relationship with the supernatural world and in such societies most diseases are believed to be caused by afflictions from supernatural forces (Alem et al., 1999; Mulatu, 1999; Ayano et al., 2015). Supernatural causes are reported to include but not limited to; demon possession, bewitchments by evil spirits, ancestor’s spirits and/or the evil eye (Ayano et al., 2015). The Tanqway (witch doctors) tend to be consulted for conditions perceived to have a supernatural cause, i.e., psychosis (Selamu et al., 2015).

Historically, anthropological studies have reported that Ethiopians attribute mental illness to both supernatural and natural causes, however, the former has been implicated as a belief most widely held amongst the Ethiopian populace (Mulatu, 1999). The claim that lay people in Ethiopia exclusively believe in supernatural causes of mental illness has been challenged by findings from Mulatu (1999), who found, in a study interviewing 455 adults in the city of Bahir Dar (located in northwestern Ethiopia) that schizophrenia was believed by many to have a strong stress-related origin (e.g., loss of a loved one, lack of employment opportunity and conflict within the family). Moreover, in a community based cross sectional study conducted in Hawassa city, Ayano and colleagues (2015) found that two-thirds of their respondents reported mental illness to be caused by either poverty, infections, use of substance, loss of loved one, conflict with family or hereditary.

Some key determinants of whether one attributes supernatural or natural causes to the development and maintenance of mental illness in Ethiopia are level of education, socioeconomic status and domicile in either a rural or urban setting (Mulatu, 1999; Solomon et al., 2018; Girma et al., 2024). In a cross-sectional study conducted among Theological college students, in which the Short Explanatory Model Interview was used to assess the perception of causes of schizophrenia, Solomon and colleagues (2018) found that 76.5% (313) of their participants attributed schizophrenia to psychosocial causes in comparison to 16.9% who attributed schizophrenia to supernatural causes. The authors posit that this finding may have resulted from their participant population being educated up to the college level. Furthermore, the authors speculate whether this finding could also be in part due to the geographical origin or domicile of their participants, as more than half their sample population were from urban settings. This postulate has also been explored by Girma and colleagues (2024), who found in a qualitative study in which 16 participants were interviewed in attempt to understand mental health-related stigma that those from less educated areas of the countryside were more likely to report that mental illness was caused by spirit possession, black magic, or astrological misalignment.

**Treatment approaches**

In Ethiopia, modern psychiatric services and mental health specialists remain scarce, ultimately impacting service provision (Bekele et al., 2009; Hanlon, 2017). Consequently, Ethiopia primarily relies on psychiatric nurses to implement and deliver treatments for psychosis (Hanlon, 2017; Alem et al., 1999). The psychiatric nurses’ training programme was started in 1986 and by 1997, 140 nurses had been trained. The training was designed to enable them to identify and treat common psychiatric disorders (Alem et al., 1999). More recently, the Ethiopian mental health care plan has shifted its focus to community-based awareness-raising and case detection, with primary care workers (nurses and health officers) tasked with making the diagnosis, initiating treatment (including prescription of psychotropic medication) and providing continuing care, with monthly supervision from a psychiatric nurse (Hanlon, 2017).

The WHO Mental Health Gap Action Programme (mhGAP) aims at scaling up services for mental, neurological and substance use disorders for countries especially with low- and middle-income. In Ethiopia both psychiatric and primary care workers receive mhGAP training and are expected to follow protocol when treating psychoses (Hanlon, 2017). The protocol outlines the first stage of management of psychoses is to provide psychoeducation to the person and caregivers. The second stage is the commencement of antipsychotic medication, starting with a low dose within therapeutic range. Next is to promote functioning in daily activities; then ensuring the safety of the person and others; and finally providing regular follow-up. If the individual is well enough, they move to receiving support and rehabilitation in the community, whilst given assistance to reduce stress and strengthen social support (WHO, 2016). Engagement with traditional and faith healers is also recommended as an essential part of community provision of care for people with SMI, but this is rarely put into practice and even more rarely evaluated (Hanlon, 2017). Although the mhGAP programme has been successfully adapted to the situation in Ethiopia by the Ministry of Health, and though these policy initiatives are necessary, it is acknowledged that more needs to be done (Fekadu & Thornicroft, 2014).

There has been some burgeoning work on psychosocial interventions for schizophrenia in Ethiopia. In a mixed methods pilot study that took place in rural Ethiopia, in which ten people with schizophrenia, unresponsive to treatment with medication alone, and their caregivers participated in Community Based Rehabilitation (CBR) trial (Asher et al., 2018), found CBR to have a positive impact. In the trial participants were given psychoeducation as part of the health component of CBR.^[2]^ This involved 30–90-minute weekly home visits by a community-based rehabilitation worker (CBRWs) who received 5 weeks of training in CBR delivery and basic counselling. The programme was reported to enhance family support, improve access to health care, boost income and improve self-esteem of participants. The authors concluded that CBR is an acceptable and feasible adjunct approach to facility-based care for people with schizophrenia. These findings demonstrate that there are benefits to be found from psychosocial interventions in Ethiopia and possibly towards the acceptability of AVATAR therapy.

Although the Ethiopian mental health care plan seeks to prioritize modern psychiatric and psychosocial treatment over traditional methods of treatment (e.g., faith healer, herbalist) (Hanlon, 2017), in practice, this may not be the case. As in a paper on the patterns of treatment seeking behaviour for mental illnesses in Southwest Ethiopia: a hospital-based study, the authors reported that traditional healers were the first place where help was sought for mental illness in their population (Girma & Tesfaye, 2011). It has been suggested one reason for this is low population coverage of specialist mental health services in Ethiopia (Hanlon et al., 2024). This seems to be further backed up by the work of Girma and Tesfaye (2011) who reported that nearly all their respondents 379 (98.7%) believed that mental illness can be cured with modern treatment, demonstrating that there is demand for more modern psychiatric treatments in Ethiopia. However, there seems to be a lack of published research in the English language on patient led outcomes (Souraya et al., 2018), with most of the published literature reporting asymmetric outcomes, centring around general course of illness, patterns of remission, and the percentage of time cases spent in different clinical states (Alem et al., 2009)

In a qualitative study conducted in rural Ethiopia as part of the Rehabilitation Intervention for people with Schizophrenia in Ethiopia (RISE) project, Souraya and colleagues (2018) reported that people with schizophrenia and caregivers were often unable to execute their desired choice due to inaccessible and unaffordable treatment. The authors concluded that further studies are needed to explore concepts of person-centred care and recovery across cultural settings (Souraya et al., 2018; Hanlon, 2017).

The main biomedical intervention for the treatment of schizophrenia in Ethiopia is the use of antipsychotic medication; however, studies are revealing high rates of non-adherence (Alem et al., 2009; Assefa et al., 2012; Souraya et al., 2018; Teferra et al., 2012, 2013) This is consistent with patterns observed in the UK, where medication is frequently prescribed but where acceptability and adherence vary. (Teferra et al., 2013) list potential reasons for non-adherence in Ethiopia, including the effects and experiences of medication being impacted by other variables (e.g., food shortages and/or insecurities); lack of social and family support; and expectations of cure, rather than medication being needed long-term. These findings may point to a greater need of alternative or adjunct treatments, and a potential role for psychosocial interventions.

**Implications for AVATAR therapy**

It is likely that there are important differences in the way that psychosis is understood, experienced, and treated in Ethiopia, compared to the UK. There is a common influence of spiritual and supernatural beliefs when it comes to understanding the experience of voice-hearing. It is only relatively recently that psychiatric and community approaches to the treatment of psychosis have been introduced in Ethiopia, and so there is a lot still to learn about the acceptability and effectiveness of these approaches. Even less is known about psychosocial interventions for psychosis and for voice-hearing specifically. Factors such as gender, age, urbanicity, educational level, and spiritual beliefs might impact on individuals’ keenness to receive psychological therapy and the acceptability of the AVATAR intervention specifically.

Published trials to date have evaluated biomedical interventions, particularly the use of antipsychotic medication (Souraya et al., 2018; Alem et al., 2009) and interventions provided by psychiatric nurses and mental health workers (Hanlon, 2017). Case studies from non-governmental organisations provide examples of holistic approaches to rehabilitation, recovery and empowerment of people with SMI (please see Asher et al., 2015; Asher et al., 2022; and Asher et al., 2018 for in depth discussion of CBR for Schizophrenia in Ethiopia), but rigorous comparative studies are needed to identify the most efficient, effective and scalable approaches to care (Hanlon, 2017). Given the barriers to mental health care highlighted above, it will be important to establish what care pathways look like for psychoses in Addis Ababa in order to fully support prospective deliverers of AVATAR therapy.

**Conclusions and questions**

It will be important to understand how individuals within the different communities of Ethiopia relate to voices, particularly whether voices are seen as persecutory or distressing; in AVATAR Therapy, the focus is always on what is distressing and interfering with life (Ward et al., 2020). The research has shown that although supernatural explanations for mental illnesses remain common within much of Ethiopian society, alternative explanations like psychosocial and biomedical causes are also held by some within society (Mulatu, 1999; Solomon et al., 2018; Girma et al., 2024). Sociodemographic and socioeconomic factors are reported to be key determinants of whether supernatural or biopsychosocial causes are believed (Mulatu, 1999; Solomon et al., 2018; Girma et al., 2024; Ayano et al., 2015).

There appears to be a dearth of published work in the English language on the phenomenology of voice hearing in Ethiopia. It will be important to understand more about how voices are experienced and embodied, and the extent to which individuals perceive themselves to be in a relationship and communicating with the voice that they hear, as this will have implications for the acceptability of, and adaptations needed to, AVATAR therapy.

1. LMIC is widely used but can be problematic (Lencucha & Neupane, 2022) – its use here reflects the limited access to psychological interventions common to both India and Ethiopia.
2. A Community based rehabilitation (CBR) programme is formed by one or more activities in one or more of the five components (health, education, livelihood, social, empowerment).
