## Supplementary File 1 for "Feasibility and acceptability of contextually adapted AVATAR therapy for distressing voices in Ethiopia and India: a study protocol for the AVATAR3 study"

**Lived Experience Framework for AVATAR3**

*At this stage, this document (v1) has been led by Sophie Ul-Haq, lived experience lead researcher based in the UK, and is subject to ongoing review by the international partners.*

**Ethos of lived experience involvement in AVATAR**

Lived experience (LE) is more than just direct personal experience, it is a form of expertise in its own right. The AVATAR team recognises that LE is a unique, person-centred form of knowledge, insight and expertise that comes from living through or with mental health challenges.^[1]^

We will involve people with LE throughout the AVATAR3 study (both International and AI work) creating a multidisciplinary team that can work creatively and collaboratively, taking basic ideas transforming them into truly impactful pieces of work.

LE colleagues will be involved in recruitment, decision-making, co-design, study design, data collection, analysis, dissemination and the evaluation of this project.

LE brings often overlooked perspectives to the field of mental health. It ensures the research conducted is pertinent to real life concerns and can be applied in practice.

People with LE have a nuanced understanding of the challenges, emotional impacts, cultural context and systems within MH services. When meaningfully integrated, this expertise functions as a vital skillset encompassing analytical, relational and strategic capacities that can drive innovation, foster empathy and support the transformation of mental health systems for the benefit of all.

By including diverse LE perspectives within research teams, new knowledge can be generated that reflects a broader range of experiences and realities. We strongly believe that integrating LE in this work is the right thing to do ethically and that we should not be making decisions without the people that are most affected by them.

In this project, LE will be valued in the same way as other forms of expertise. LE colleagues are encouraged to bring their whole selves to this work as they will have a range of skills, understanding and perspectives that will advance this work and take it in new directions.

Reflecting on practice around LE involvement in informal debriefs or supervisory spaces ensures that this work is not only meaningful but also continually evolving.

Capturing the learning emerging from LE involvement and the impact of this is also a key part of this project.

Meaningful and diverse LE involvement will not only enhance trust in the work undertaken but also serve as a key element in ensuring the study is conducted ethically and responsibly.

**Commitments to Lived Experience Colleagues**

- People with LE are involved from the earliest stages, when ideas are still forming and meaningful change is possible.
- Honesty, transparency, and openness will guide all interactions
- Materials, meetings and requests are made clear and accessible to everyone.
- Involvement can be flexible and tailored to the person
- Providing the right support to facilitate this, recognising that capacity to take part may fluctuate for many reasons
- Decision-making processes are clear, fair and easy to understand.
- A respectful environment is created where every contribution is valued and everyone feels empowered to shape the work.
- Each person is recognised as an individual — with unique hopes, strengths, and challenges — and approaches are shaped together to reflect this.
- The offer of reasonable adjustments for each person along with access to personal development opportunities, including training and supervision (for example around involvement in qualitative aspects of study) as well as information on wider LE opportunities outside of the project.

**Power imbalances.** Within this work we need to consider and address power imbalances between those with LE, clinicians, researchers, and other mental health professionals but also computer programmers, designers etc. Acknowledging these power imbalances ensures that the research is ethical and respectful, mitigating the risk of harm. This acknowledgement also allows us to implement strategies that minimise these power imbalances thereby enhancing the validity and reliability of the work conducted.

**What lived experience colleagues contribute to the research:**

- Provide authentic, real-world insights that make research more relevant and grounded.
- Highlight issues and priorities that may otherwise be overlooked.
- Help ensure research questions, methods, and outcomes reflect real needs and experiences.
- Contribute to ethical sensitivity by identifying potential risks or harms in design or implementation.
- Help identify anything that may make participants in the research feel uncomfortable
- Improve the accessibility and clarity of study materials (e.g., participant information sheets).
- Enhance trust and engagement between researchers and participant communities.
- Strengthen dissemination by helping to interpret findings in meaningful, relatable ways.
- Challenge assumptions and biases within the research team.
- Promote inclusivity and diversity in both participation and interpretation.

Excellent LE involvement within this project is characterized by the active and meaningful participation of diverse voices, ensuring that individuals from various backgrounds, including those from marginalised or underrepresented communities are instrumental in shaping the direction and outcomes of the work. This involvement goes beyond consultation to tangible changes that improve the quality, accessibility, and ethical integrity of the project. In the specific contexts of India and Ethiopia, the work is adapted in culturally sensitive and contextually relevant ways, ensuring that local norms, values and lived realities are reflected in the research process and outcomes. LE colleagues are empowered to gain new skills, build confidence, and acquire valuable experiences that extend beyond the project itself, contributing to their personal growth and broader community impact. We commit to working relationally, fostering deep, trust-based connections with participants, and providing tailored support and flexibility so that each individual can engage in the project in a way that suits their needs, preferences, and personal circumstances. Ultimately, excellent LE involvement is about creating a space where everyone’s input is valued, where people feel respected and supported, and where their contributions lead to meaningful and lasting change.

**What lived experience colleagues can gain from being involved:**

- Enjoyment and finding acceptance
- A sense of empowerment and validation through having their experiences valued.
- Opportunities to influence change and improve services or policies.
- Development of new skills (e.g. communication, research literacy, data interpretation).
- Greater understanding of research processes and evidence-based practice.
- Confidence, increased self-esteem, and pride in contributing to knowledge creation.
- Connection and shared purpose with other contributors and researchers.
- Potential career or volunteer pathways in research, advocacy, or peer support.
- Personal growth through reflection on their own experiences

**Support Offered to Lived Experience Colleagues**

**Induction materials and support for new members of the lived experience team:**

- 1-2-1 meeting with Patient and Public Involvement (PPI) lead researcher
- An overview to the project including the history, context and aims is shared so everyone understands the bigger picture
- Information about the project is provided clearly, ensuring understanding of what is expected at each stage
- A named Research Assistant (RA) acts as a main point of contact
- Identifying any accessibility requirements or support needed and the AVATAR team putting this support into place
- Introduction to the project via various resources
- Setting up payment – Lived Experience lead researcher discusses payment options and helps identify the most suitable approach and sets this up.
- Agreeing availability and how the person would like to be involved – offering flexibility within this

**Emotional/ Wellbeing**

**Processes in creating a ‘safe enough’ space^[2]^:**

- Acknowledging the emotional labour and the potential impact of being involved in such a role - providing the time and space to explore this, if desired.
- Creating a non-judgemental and supportive atmosphere when facilitating meetings
- Allow people to bring their whole selves to the work – encourage them to ask questions, voice concerns and challenges, and engage in ways that feel authentic to them
- Lived Experience Lead researcher using their own experiences to enable others to feel comfortable to share while not centring these
- Building relationships with people where they feel able to come to the team at any point during the work
- Allowing people to have the space to not have to directly name own their experiences if they feel more comfortable and teaching them how to do this so that they feel less exposed
- Sending out advance notice if the content of the discussion/ documents/ videos etc. may be distressing or unexpected
- Option of Mentoring/ Buddying by the Research Assistants (RA’s)
- Briefing and debriefing of individuals before and after sessions
- Recognizing and respecting the diversity of experiences in the group, including around race, gender, socioeconomic status, and other lived experiences.

**Accessibility**

- Meetings are designed to be accessible — in format (digital and in-person), venue and in the way they are structured.
- Tech support will be offered along with test runs for online attendees to avoid digital exclusion.
- Resources are adapted to ensure clarity and accessibility for all.
- LE colleagues can opt in or out of parts of the meeting (especially if sensitive topics are discussed).
- A relaxed and welcoming atmosphere is created, with space for icebreakers and unstructured conversation.
- We will offer regular breaks within the sessions and the opportunity for a debrief afterwards.
- Opportunities are created to build skills and confidence through active participation.
- Multiple ways of contributing are offered, including in-person, online, and offline options.
- The Lived Experience lead will maintain communication between meetings, valuing LE as an ongoing partnership.
- LE colleagues will be asked how the meeting felt for them and how accessibility could improve next time.

**Support and Resources**

Any support that is offered or put in place should be discussed with and actioned in collaboration with that individual. We recognise that those with LE are experts in their own experience and are often best placed to tell us exactly what they need.

**Potential sources of support in crisis:**

SLAM mental health Crisis line: 0800 731 2864

[NHS 111 online](https://111.nhs.uk/triage/check-your-mental-health-symptoms)

We will also offer all those involved from a LE perspective the option of creating a personalised plan around what they would like to happen if they are working for us and struggling with their mental health.

**Remuneration**

As part of ensuring that LE in valued in same way as other forms of expertise it is vital that those with LE are paid for their time and involvement in the project (following the established AVATAR3 payment policy).

**Recognition for Lived Experience Colleagues**

There will be multiple opportunities for LE colleagues to be recognised for their contributions to the research, this may be in several formats such as:

- Presentations by the research team and LE colleagues
- Open Science Framework Website – sharing the plans for the research
- Conferences
- Academic Publications
- Public-facing media e.g. website, social media posts.

The research team will always discuss with individual LE colleagues how they would like to be recognised for their input, there will be several options including:

- Full name
- First name only
- Alias
- Consortium e.g. the AVATAR3 PPI Group

The preference indicated by the PPI member will be followed by the research team and can be changed at any time.

**An Overview of the AVATAR3 Project**

AVATAR therapy is a targeted intervention aimed at reducing the distress associated with auditory verbal hallucinations (voices). In the UK work, the aim is to test automation of avatar dialogues, which represent a core component of each therapy session, using Artificial Intelligence (AI) powered conversational agents. By demonstrating that it is possible to train a widely available UK workforce to support AVATAR AI, we will deliver a step-change in UK-based implementation of AVATAR therapy. The AVATAR3 project also aims to test the acceptability and feasibility of AVATAR therapy delivered across two diverse Low- and Middle-Income countries (LMIC) India and Ethiopia. The vision for the project is to deliver AVATAR therapy as an effective, globally scalable intervention for people experiencing distressing voices. Our focus is on national and international implementation of AVATAR therapy with the collection of outcome data in real-world contexts.

**Proposed Governance Structure for Lived Experience Involvement**

We are aiming to actively recruit and involve a broad range of people with demographic diversity in this work, including members of marginalised groups and people who may be sceptical about the research.

**Diagram of proposed governance structure and how this links to project team:**

**Lived Experience Lead Researcher**

**Mental Health Service User Association (MHSUA) - Ethiopia**

**AVATAR AI Lived Co-design group**

**Indian Lived Experience Lead Researcher**

**Planned consultations**

**Planned consultations**

**Lived experience advisory group**

**Planned consultations**

**User experience testing**

*The purpose of this structure is to set a clear intention with respect to the involvement of LE colleagues. We recognise that such an approach is necessary to allow individuals with lived experience to thrive, have an impact within the work and meaningfully engage in decision-making.*

**Overview of Different Roles within the Governance Structure**

**Lived Experience Lead Researcher.** This role involves collaboration and strategic oversight across both workstreams of the project. In addition to engaging in the day-to-day operational aspects, the Lived Experience Lead researcher will provide high-level oversight and guidance, offering critical insight into the overall direction of the project to ensure that the principles of meaningful and ethical LE involvement, as well as alignment with LE values and priorities, are embedded throughout the research process. They will facilitate the AVATAR AI co-design group – ensuring these meetings are accessible and meaningful. Their role also involves providing support to those in LE leadership roles in Ethiopia and India. The Lived Experience Lead Researcher will collaborate with international partners to build and develop Expert by Experience (EbE) models that are tailored to the context in which they are situated. Their role will be to monitor and assess involvement in both workstreams, identify additional opportunities for those with LE and to brainstorm around where this involvement could be strengthened. They will create a space where challenges in practice can be discussed openly and creatively and where experience-informed solutions can be developed. The LE lead researcher’s role offering accountability, vision, and constructive challenge to support the meaningful transformation of mental health research practices. They will also build individual relationships with the PPI consultants to allow for direct feedback on the work and processes around this.

**AVATAR AI Work**

**AVATAR AI Co-Design Working Group – Proposed role.** The AVATAR AI Co-Design Working Group will bring together a small but diverse group of people with LE including those from marginalised and underrepresented communities and those that are sceptical about the use of AI. The group will play an active and collaborative role in shaping the development and ethical use of AVATAR-based AI technologies within the project. Their role is to ensure the co-design process is grounded in real-world relevance and responsive to a range of perspectives. The voices of this group will directly influence how accessibility, acceptability and trust are designed into the system from the outset. We will offer a space where new insights from those often overlooked by traditional innovation processes can guide more inclusive development. This group will also focus on user acceptability looking at how the AVATAR AI therapy is framed, assess realism and determine usability.

We will also engage a broader group of people around core aspects of the design to ensure diverse perspectives are represented, address any gaps, and explore specific issues that may arise through the course of the work.

**User Experience Testing of AVATAR AI.** We will undertake user testing with a broad and diverse group of patients and carers, intentionally including individuals from marginalised and underrepresented communities. Importantly, those involved in the user testing will have had no prior involvement with the project or exposure to the co-design process. This ensures that their responses to the AVATAR AI are fresh, uninfluenced and reflective of authentic first-time user experiences—similar to how future clients might encounter the therapy in real-world settings. Their feedback will play a critical role in evaluating the accessibility, relevance and emotional resonance of the intervention as well as how realistic the AI AVATAR is perceived to be.

**AVATAR International Work**

The Ethiopian and Indian site will share learning and establish an involvement legacy, tailored to each distinct context. We will work with individuals from a diverse range of cultural, spiritual, socio-economic, gender and linguistic backgrounds to inform the development of AVATAR International. LE at each site will provide critical insights into how the therapy is likely to be perceived, received and experienced across different contexts, helping to ensure cultural sensitivity, relevance, and ethical integrity. Each group will contribute to co-developing inclusive, person-centred approaches that reflect the values and needs of varied communities.

In Ethiopia, LE involvement will be led by the **Mental Health Service User Association of Ethiopia (MHSUA)** as a research partner. MHSUA-Ethiopia has been a collaborator from the inception of the project, including the development of the research proposal. While MHSUA have previously collaborated on a range of research projects, this study is the first time the organization has been formally contracted within the collaborator agreements, meaning the organization holds a budget linked to work they will lead in Ethiopia and an MHSUA employed Research Worker. This represents a crucial step in capacity building and equity in partnership and helps to ensure that LE perspectives remain central throughout the study. Ongoing MHSUA involvement and leadership include full participation at project site meetings, an MHSUA representative leading the LE discussion at the UK Responsible Research and Innovation (RRI) event, involvement in cultural adaptation both through co-production of the tools contributions to ongoing adaptation planning, co-production of materials designed to support participation, involvement in inclusive stakeholder engagement (both co-production of the inclusive stakeholder engagement methods and supporting participation of members at community events).

At the **All-India Institute of Medical Science (AIIMS)** the team will build on successful LE involvement and co-design during a previous project, which developed a blended digital intervention (Saksham) for people with a diagnosis of schizophrenia (and caregivers). The AIIMS team will recruit to a LE lead role who will receive training to co-deliver qualitative interviews and be involved in analysis. The team will also form a wider LE group comprising individuals with LE of voice-hearing and their carers who will be involved across diverse research activities.

**Pivotal Points for Involving those with Lived Experience in the AVATAR Project**

It is vitally important that we define the essential points at which people who hear voices and their carers are involved in the project from the outset. This helps us to be accountable to these and ensure that things are less likely to be overlooked. These pivotal points include but are not limited to:

- **Prioritisation:** Identifying research priorities, key outcomes, and way of modifying the approach to research questions. Supporting alignment of research aims with LE perspectives to ensure relevance and impact.
- **Design and Development:** Co-developing inclusive, empowering, and non-stigmatising solutions. Advising on ethical issues, feasibility, and practical challenges. Ensuring marginalised voices are represented and contributing to iterative design of AVATAR therapy, including interface, language, and user experience.
- **Governance and Leadership:** Contributing to co-design groups, strategic discussions and leading the vision around LE involvement, ensuring discussions are grounded in real world experiences.
- **Data Collection:** Designing and reviewing data collection tools and methods. Taking part in qualitative interviews and helping to ensure data collection reflects diverse perspectives.
- **Analysis and Interpretation:** Collaborating on the evaluation of research findings and exploring their meaning, implications, and potential for impact through a LE lens.
- **Reporting and Dissemination:** Co-creating academic and public-facing outputs such as papers, blogs, and presentations. Supporting strategies for effective dissemination to research communities, LE networks, and the wider public.
- **Evaluation:** Helping to reflect on how knowledge is shared and used. Evaluating LE shapes how the project is carried out and what is learned along the way.

**Methods for Evaluating Lived Experience Engagement**

Methods for evaluating LE engagement will be built in from the start by capturing the points at which LE makes meaningful changes within the project, ensuring that the impact of this involvement is visible and valued throughout. This will be supported by a co-developed evaluation framework, created with LE contributors, beginning with a shared understanding of what meaningful involvement looks and feels like, including which outcomes matter most to those with LE.

We will set clear indicators and review points at key stages of the project and will document how the project evolves in response to LE input. Regular, informal drop-ins combining both structured and unstructured elements will allow us to gather rich, ongoing feedback on contributors’ experiences and perceptions.

We will ensure findings are shared regularly with the entire team, including those with LE and make the insights clear and actionable.

By embedding these reflective and responsive processes from the outset, we will ensure LE involvement is not only meaningful but continuously evaluated and strengthened across the life of the project.

1. Wellcome (n.d.) *Embedding lived experience expertise in mental health research.* Available at: <https://wellcome.org/research-funding/guidance/prepare-to-apply/embedding-lived-experience-expertise-mental-health-research> (Accessed 15^th^ December 2025).
2. Mind (2024). *Mind Lived Experience Framework.* Available at: <https://www.mindaustralia.org.au/sites/default/files/2024-09/Mind_Lived_Experience_Framework.pdf> (Accessed 15^th^ December 2025).
